# Longitudinal insights into dynamic patterns and cumulative burdens of biological age acceleration in relation to type II diabetes mellitus, all-cause mortality and glycemic traits

**DOI:** 10.64898/2026.04.07.26350301

**Authors:** Yu Yan, Chu Zheng, Ping Zeng

## Abstract

**Background:** Accelerated biological aging (BioAgeAccel) has been implicated in type II diabetes (T2D) mellitus development; however, its dynamic changes and their links to T2D incidence, mortality and glycemic traits remain unclear.

**Methods:** Leveraging repeated measures from the UK Biobank, we first calculated two BioAgeAccel metrics (KDMAccel and PhenoAgeAccel) and derived three burdens (slope, cumulative, and relative cumulative change). We then assessed associations of BioAgeAccel transitions and these burdens with incident T2D and mortality. Secondary analyses extended the two primary outcomes by incorporating glucose, HbA1c, and six IR surrogates, which were also evaluated as potential mediators.

**Results:** Among 13,751 included participants, 412 (3.0%) new T2D cases and 609 (4.4%) all-cause deaths were identified within a median follow-up of 9.5 years. Dynamic transition from non-accelerated to accelerated aging was markedly related to elevated T2D risk (KDMAccel: HR=1.65 [1.24∼2.20]; PhenoAgeAccel: HR=1.50 [1.12∼2.00]) and all-cause mortality risk (KDMAccel: HR=1.32 [1.06∼1.64]; PhenoAgeAccel: HR=2.17 [1.73∼2.71]). BioAgeAccel burdens demonstrated dose-response effects, with cumulative BioAgeAccel showing the greatest influence on T2D (KDMAccel: HR=1.25 [1.03∼1.51]; PhenoAgeAccel: HR=1.26 [1.06∼1.49]) and all-cause mortality (KDMAccel: HR=1.25 [1.07∼1.47]; PhenoAgeAccel: HR=1.51 [1.31∼1.74]). Similar association patterns were observed for all the eight glycemic traits. Mediation analyses revealed that these glycemic traits on average mediated 19∼32% of the KDMAccel burden-T2D effect and 16∼24% of the PhenoAgeAccel burden-T2D effect. Incorporating BioAgeAccel burden into FINDRISC significantly enhanced prediction accuracy, reaching up to 10.9% improvement in some specific aging transition statuses.

**Conclusion:** Dynamic biological aging trajectories and BioAgeAccel burdens are independently related to elevated risks of T2D and all-cause mortality, partly via glycemic dysregulation, highlighting biological aging as a potential intervention target.

## Introduction

Type II diabetes (T2D) mellitus is a complex metabolic disorder characterized by impaired glucose metabolism, primarily driven by insulin resistance (IR) and β-cell dysfunction (DeFronzo, et al., 2015). Over recent decades, the global prevalence of T2D has increased dramatically, posing a significant public health challenge (Ong, et al., 2023). While traditional risk factors including obesity (Li, et al., 2021), sedentary lifestyle (Kolb and Martin, 2017), and genetic predisposition (Suzuki, et al., 2024) are well-established, growing evidence implicates biological aging — the progressive decline in physiological function — as a critical contributor to T2D pathogenesis (Bahour, et al., 2022; Jiang, et al., 2024; Pan, et al., 2025; Zhang, et al., 2025). Therefore, elucidating the interplay between biological aging and T2D may reveal novel insights into mechanisms underlying glycemic dysfunction and inform targeted interventions.

Although chronological age remains a primary determinant of T2D risk, it fails to account for interindividual variability in aging trajectories, where individuals of identical age exhibit markedly different metabolic and physiological decline (Ahadi, et al., 2020). To address this limitation, biological age (BioAge) and its acceleration (BioAgeAccel) have emerged as more accurate metrics for quantifying aging-related physiological dysregulation or deterioration (Jylhävä, et al., 2017). Among these, Klemera-Doubal method biological age (KDM) and phenotypic age (PhenoAge) are two well-validated algorithms that leverage different sets of clinical markers such as blood chemistry profiles, lung function, and blood pressure to assess aging status beyond chronological age (Klemera and Doubal, 2006; Kuo, et al., 2021; Levine, 2013; Liu, et al., 2018). Interestingly, a growing body of research has revealed that biological aging is dynamic, with evidence suggesting it can even be reversible (Chen, et al., 2024; Poganik, et al., 2023).

Although several studies have indicated that individuals with accelerated biological aging encounter higher risk of T2D (Bahour, et al., 2022; Jiang, et al., 2024; Pan, et al., 2025; Yang, et al., 2025; Zhang, et al., 2025), the dynamic link of aging with T2D risk has not yet been fully explored, partly due to lack of large-scale longitudinal data with repeated measurements. Beyond T2D, accumulating evidence has also underscored the prognostic value of BioAge in predicting mortality (He, et al., 2024; Jiang, et al., 2024; Tian, et al., 2025; Xiang, et al., 2024), independent of traditional risk factors. However, these findings were mainly driven from single-timepoint assessments, offering limited insight into the dynamic evolution of aging trajectories and their impact on metabolic diseases or death.

Further, previous work has focused primarily on the relation of aging with binary T2D outcomes rarely considering the continuum of glycemic dysregulation, including blood glucose (Tabák, et al., 2012), hemoglobin A1c (HbA1c) (International Expert Committee, 2009), and IR (Brownlee, 2005), which are pivotal for understanding early metabolic impairments and monitoring precursor symptoms of T2D. Therefore, investigating how BioAgeAccel affects this spectrum of glycemic dysregulation can unveil stage-specific mechanisms in diabetes development and provide opportunities for early intervention. Moreover, no studies have examined the predictive capacity of BioAgeAccel and its burdens across different transition statuses, which may offer deeper insights into how dynamic aging trajectories influence T2D risk.

To bridge these gaps, we here conducted a large-scale longitudinal study using data from the UK Biobank (Sudlow, et al., 2015). We aimed to: (i) investigate the impact of visit-to-visit changes in BioAgeAccel on T2D risk and all-cause mortality by categorizing participants into different transition statuses based on the stability or progression of BioAgeAccel; (ii) evaluate the influence of three BioAgeAccel burden indicators — BioAgeAccel slope, cumulative BioAgeAccel, and relative cumulative BioAgeAccel — on the risk of T2D incidence and all-cause mortality; (iii) extend the primary analysis of T2D by incorporating glucose, HbA1c, and 6 IR surrogates as secondary outcomes and mediators to provide a more comprehensive understanding of metabolic mechanisms linking BioAgeAccel to T2D risk; (v) explore whether BioAgeAccel burden could enhance conventional risk prediction models such as Finnish Diabetes Risk Score (FINDRISC) (Lindström and Tuomilehto, 2003; Peng, et al., 2023), which is a widely-employed prediction tool for identifying people with high risk of T2D and glucose impairment. The flowchart of participant enrollment and data analysis is given in Figure 1.

**Figure 1.**
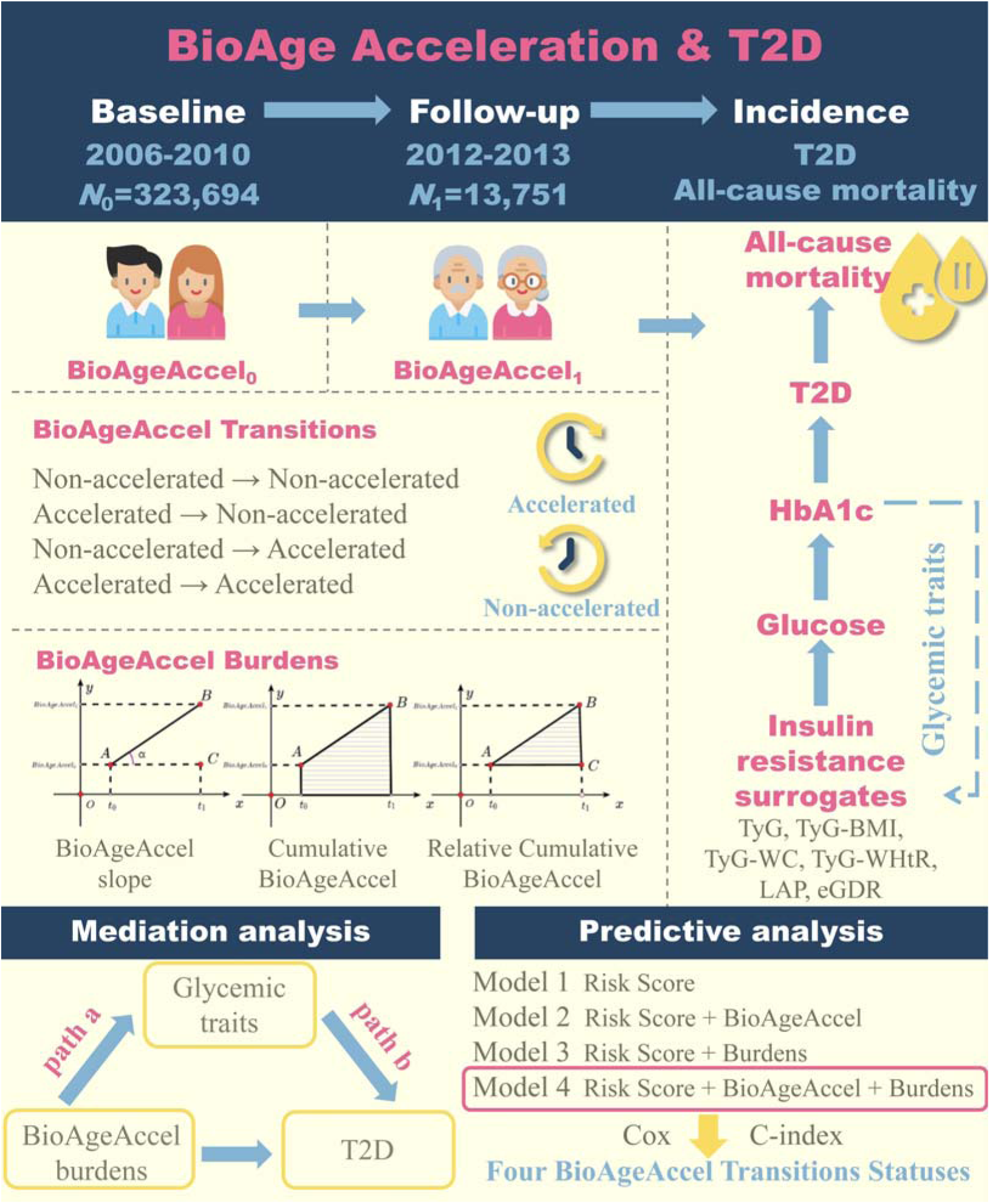
Flowchart of participant enrollment and data analysis in this study. BioAgeAccel: biological aging acceleration; BioAgeAccel_0_ and BioAgeAccel_1_ referred to BioAgeAccel for each participant at baseline and the first follow-up, respectively; T2D: type II diabetes; TyG: triglyceride-glucose; TyG-BMI: TyG combined with body mass index (BMI); TyG-WC: TyG combined with waist circumference; TyG-WHtR: TyG combined with waist-to-height ratio; LAP: lipid accumulation product; eGDR: estimated glucose disposal rate.

## Methods and Materials

### Data sources and study design

Extensive baseline data of the UK Biobank were collected between 2006 and 2010 (Sudlow, et al., 2015). During its first follow-up phase conducted from 2012 to 2013, over 20,000 participants underwent repeated assessments (Figure 1). We retained participants who completed both assessment visits and provided the necessary marker data for BioAgeAccel (*N*=13,751), while excluding those diagnosed with T2D between baseline and the first follow-up (Figure S1).

### Definitions and ascertainment of T2D, glycemic traits, and BioAge

#### Ascertainment of T2D and follow-up time

T2D was mainly ascertained via ICD-10 codes (Table S1), and some other records were also applied, such as self-reported illness (Field 20002), medical conditions (Fields 6153 and 6177). The endpoint was defined as the date of diagnosis of T2D events, all-cause death, failure of follow-up, or follow-up deadline (31 Dec 2021), whichever occurred first. The follow-up time for each participant was calculated from the first follow-up date to the endpoint.

#### Definition of glucose, HbA1c, and IR surrogates

Clinically, blood glucose, HbA1c, and IR indices represent distinct but interconnected stages along the glycemic dysregulation continuum leading to T2D. IR is often the earliest detectable abnormality, manifesting before any measurable increase in blood glucose levels. As IR worsens, glucose and HbA1c levels gradually rise, eventually surpassing diagnostic thresholds for T2D (Brownlee, 2005; International Expert Committee, 2009; Tabák, et al., 2012). Therefore, we additionally considered 6 surrogates for IR (Supplementary File), including TyG (Ramdas Nayak, et al., 2022), TyG-BMI (Er, et al., 2016), TyG-WC (Zheng, et al., 2016), TyG-WHtR (Lim, et al., 2019), LAP (Kahn, 2005), and eGDR (Zhang, et al., 2024). All glycemic traits were standardized before formal analysis.

#### Selection of confounding covariates

A wide range of sociodemographic characteristics and lifestyle factors were identified as covariates, including age, sex, education (college degree or without college degree), income (<£31,000 or ≥£31,000) (Office for National Statistics, 2017), Townsend deprivation index (TDI), drinking status (yes or no), smoking status (yes or no), healthy diet score (calculated based on daily dietary factors, with scores ranging from 0 to 5) (Wang, et al., 2021), physical activity (low [<600 MET-min/week], moderate [600-3000 MET-min/week], and high [>3000 MET-min/week]) (Boonpor, et al., 2023), and BMI. Some well-known risk factors such as family history of T2D were also included. Multiple imputation was performed via the multivariate imputation by chained equation (MICE) method to impute missing values for each covariate (van Buuren and Groothuis-Oudshoorn, 2011).

#### Calculation of biological aging acceleration

BioAge was assessed via KDM and PhenoAge — two distinct but widely recognized algorithms (Kuo, et al., 2021). Participants’ PhenoAge was calculated from nine blood chemistries and chronological age, KDM was computed using forced expiratory volume in one second, systolic blood pressure, and seven blood chemistry chemistries (Supplementary File). BioAgeAccel (KDMAccel and PhenoAgeAccel) was defined as the residual of BioAge adjusted for chronological age through a general linear model, with higher values indicating faster biological aging relative to peers. KDMAccel and PhenoAgeAccel exhibited only weak correlations (Figure S2), indicating that they capture distinct aspects of biological aging.

#### Definition of BioAgeAccel transitions and burdens

According to the BioAgeAccel status of individuals at baseline and the first follow-up, BioAgeAccel transitions were categorized as stable accelerated aging, accelerated to non-accelerated aging, non-accelerated to accelerated aging, and stable non-accelerated aging. Let BioAgeAccel_0_ (KDMAccel_0_ and PhenoAgeAccel_0_) define the measurement at baseline (2006∼2010), and BioAgeAccel_1_ (KDMAccel_1_ and PhenoAgeAccel_1_) define the measurement at the first follow-up (2012∼2013); to reflect BioAgeAccel burden, three indicators were constructed (Cui, et al., 2022; Xiang, et al., 2024; Zhang, et al., 2021)

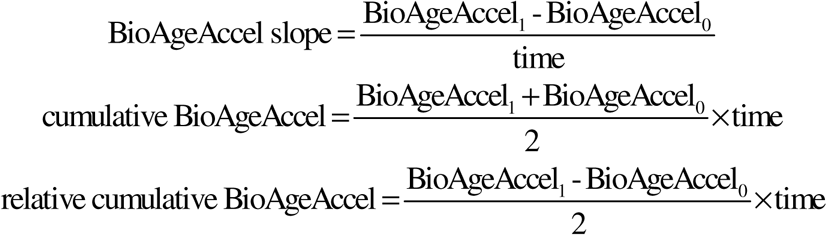

where BioAgeAccel slope stands for the rate of BioAgeAccel change, cumulative BioAgeAccel indicates the area under BioAgeAccel vs. time trajectory, relative cumulative BioAgeAccel shows cumulative BioAgeAccel that has increased or decreased relative to BioAgeAccel_0_, and time (years) is the interval between two assessments.

### Statistical methods and data analysis

#### Assess the association of BioAgeAccel with incident T2D and all-cause mortality

We employed Cox proportional hazards (PH) models to examine the association of BioAgeAccel transitions with the risk of incident T2D and all-cause mortality. No evidence regarding the violation of the PH assumption was observed according to Schoenfeld′s residuals (*P*>0.05). We here established two reference groups — stable accelerated aging and stable non-accelerated aging — to better explain the impact of BioAgeAccel progression and reversal on T2D and all-cause mortality risk. The covariates mentioned above were adjusted for when analyzing BioAgeAccel transition; but when analyzing BioAgeAccel burden, BioAgeAccel_0_ was further controlled to evaluate its independent role (He, et al., 2024; Sun, et al., 2024). Hazard ratio (HR) and its 95% confidence intervals (CI) were reported.

In addition to T2D and all-cause mortality as the primary outcomes, we also incorporated glucose, HbA1c, and 6 IR surrogates as the secondary outcomes and mediators, which provided insights into glycemic dysregulation and helped strengthen our understanding of the relation between BioAgeAccel and T2D risk. Subsequent analyses categorized BioAgeAccel burdens into tertiles (T1, T2, and T3), with the first tertile (T1) as the reference.

#### Sensitivity analyses

To validate the robustness of our results, we conducted three sensitivity analyses: (i) to minimize the reverse causality, we excluded participants who developed T2D or died during the first two years of follow-up; (ii) to evaluate the impact of drug treatments on our results, we excluded participants who underwent insulin or metformin treatments between baseline and the first follow-up; (iii) to address potential bias from competing risk (Andersen, et al., 2012), we applied cause-specific hazard models to assess the association between BioAgeAccel transitions and the risk of T2D by treating non-T2D deaths as censoring events.

#### Mediation analysis of glycemic traits in the associations between BioAgeAccel and T2D

To explore the potential mediating role of these glycemic traits in the link between BioAgeAccel and incident T2D, we here conducted a mediation analysis (VanderWeele and Vansteelandt, 2014). The relation between BioAgeAccel burden and each glycemic trait (path *a*) was evaluated via linear regression, and the link between very glycemic trait and T2D risk (path *b*) was assessed via Cox regression while adjusting for BioAgeAccel burden. A mediation effect was deemed statistically significant only when both paths yielded *P*-values<0.05 under the joint significance test framework (Zeng, et al., 2021) — a conservative criterion that minimizes false positive mediation claim. For significant glycemic traits, we estimated the mediation proportion to quantify their contributions to the total effect (Valeri and VanderWeele, 2013).

#### Evaluation of predictive capacity of BioAgeAccel to FINDRISC

Finally, we examined whether the inclusion of BioAgeAccel and BioAgeAccel burdens into FINDRISC could improve the predictive capacity (Lindström and Tuomilehto, 2003; Peng, et al., 2023). We constructed four models: (i) baseline model with only FINDRISC; (ii) model incorporating FINDRISC, and BioAgeAccel; (iii) model with FINDRISC and all BioAgeAccel burdens; and (iv) model including FINDRISC, BioAgeAccel, and all BioAgeAccel burdens. The prediction performance was validated through internal ten-fold cross-validation and evaluated via C-index. To further explore the predictive performance of BioAgeAccel burdens across distinct aging trajectories, we conducted the prediction by the four transition statuses. This enabled us to evaluate whether the predictive capacity of BioAgeAccel burdens relied on individuals’ aging trajectory. For each transition status, we calculated the C-index using the optimal prediction model.

#### Statistical software and packages

All analyses were conducted within the R (version 4.3.0) statistical computing environment. The R mice package was applied for missing value imputation (van Buuren and Groothuis-Oudshoorn, 2011), the R BioAge package was employed to compute BioAgeAccel. Two-tailed *P*-value<0.05 was considered statistically significant. The mediation analysis was conducted using the R CMAverse package (Shi, et al., 2021).

## Results

### Baseline characteristics

#### Dynamic BioAgeAccel transitions between two assessments

After the first follow-up of the UK Biobank cohort, among 13,751 participants, 412 (3.0%) new T2D cases and 609 (4.4%) all-cause deaths were identified within a median follow-up of 9.5 years (interquartile range [IQR]=9.2∼9.7). Increased BioAgeAccel was observed at the first follow-up compared with baseline (Figure S3); on average, KDMAccel elevated from -1.3 (±2.5) at baseline to -0.3 (±2.7) at the first follow-up, whereas PhenoAgeAccel rose from -6.7 (±3.8) to -5.1 (±3.9).

During a median follow-up interval of 4.9 years (IQR=2.8∼6.8) between the baseline and the first follow-up, many participants maintained their baseline aging status: 6,744 (49.0%) (KDMAccel) and 12,052 (87.6%) (PhenoAgeAccel) remained stable non-accelerated, while 3,130 (22.8%) and 325 (2.4%) respectively maintained accelerated aging. The transition patterns differed between KDMAccel (952 [6.9%] progression vs. 2925 [21.3%] reversal) and PhenoAgeAccel (363 [2.6%] progression vs. 1011 [7.4%] reversal) (Tables S2-S3).

Further, significant differences existed in the cumulative risk of T2D and all-cause mortality among various transition statuses of KDMAccel (*P*=4.24×10^-25^ for T2D, *P*=1.63×10^-4^ for all-cause mortality) and PhenoAgeAccel (*P*=2.30×10^-21^ for T2D, *P*=3.71×10^-39^ for all-cause mortality) (Figures S4-S5). Again, here slight different patterns of cumulative risk were observed for some transitions of the two BioAgeAccel measures.

#### Comparison of characteristics in different BioAgeAccel transitions

As detailed in Tables S2-S3, participants who progressed to or maintained accelerated aging consistently demonstrated less favorable baseline profiles across metabolic, socioeconomic, and lifestyle factors compared with stable non-accelerated individuals. Those reversing acceleration showed intermediate characteristics, suggesting partial recovery potential. PhenoAgeAccel transitions revealed particularly strong gradients, with accelerated groups showing older age, male predominance, and more adverse health behaviors and socioeconomic indicators.

### Association of BioAgeAccel transitions with incident T2D, all-cause mortality, glucose, HbA1c and 6 IR surrogates

#### Association of BioAgeAccel transitions with incident T2D

We observed a clear gradient in the risk of incident T2D across different BioAgeAccel transition statuses (Figure 2A). For per SD increment in KDMAccel, participants who changed from non-accelerated to accelerated aging exhibited a 65.0% (23.8∼120.0%) higher risk compared with those with stable non-accelerated aging. Participants who maintained in the stable accelerated aging status experienced an even greater risk increase of 151.0% (94.7∼223.5%). Notably, individuals who transitioned from accelerated to non-accelerated aging showed a 107.1% (45.4∼194.9%) greater risk compared with those keeping stable non-accelerated aging, suggesting that prior accelerated aging likely exerted lasting effects on T2D risk. In contrast, participants who transitioned from accelerated to non-accelerated aging demonstrated a reduced T2D risk compared with those maintaining stable accelerated aging (hazard ratio [HR]=0.83), although such reduction was insignificant (*P*=0.267). PhenoAgeAccel showed analogous results but displayed relatively attenuated influences.

**Figure 2.**
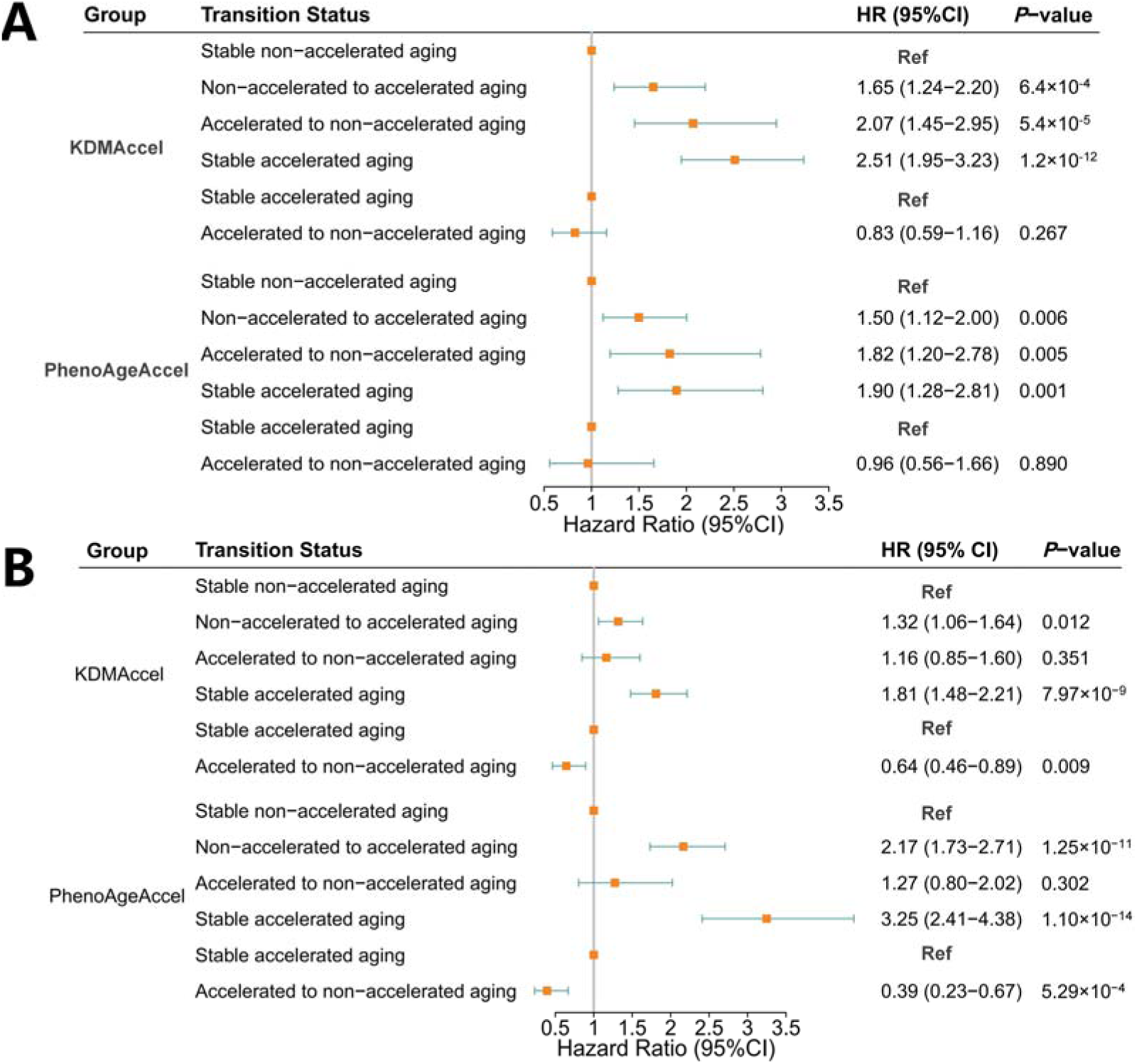
(A) Forest plots of the association of BioAgeAccel transitions with incident T2D; (B) Forest plots of the association of BioAgeAccel transitions with all-cause mortality. BioAgeAccel: biological aging acceleration; KDMAccel: Klemera-Doubal method biological age acceleration; PhenoAgeAccel: phenotypic age acceleration; T2D: type II diabetes.

#### Association of BioAgeAccel transitions with all-cause mortality

Similarly to T2D incidence, analogous association patterns were detected for all-cause mortality (Figure 2B). For per SD increase in KDMAccel, participants who transitioned from non-accelerated to accelerated aging exhibited a 31.8% (6.2∼63.6%) higher risk of death compared with those with stable non-accelerated aging, and those remaining in stable accelerated aging experienced the greatest elevation in all-cause mortality risk, with an 80.9% (47.9∼121.3%) higher hazard. Interestingly, individuals who reverted from accelerated to non-accelerated aging still showed a moderately elevated mortality risk (HRL=L1.16 [0.85∼1.60]), though not statistically significant (*P*L=L0.351). When using the stable accelerated aging group as the reference, participants transitioning to non-accelerated aging demonstrated a significantly reduced all-cause mortality risk (HRL=L0.64 [0.46∼0.89]), suggesting partial but incomplete reversal of long-term all-cause mortality hazard.

PhenoAgeAccel displayed consistent but stronger link patterns. Briefly, individuals with stable accelerated aging had a markedly elevated all-cause mortality risk (HRL=L3.25 [2.41∼4.38]), and those transitioning into acceleration also showed significantly greater risk (HRL=L2.17 [1.73∼2.71]). Particularly, reversal of acceleration was associated with a substantial reduction in risk relative to persistent acceleration (HRL=L0.39 [0.23∼0.67]).

#### Association of BioAgeAccel transitions with glucose, HbA1c and 6 IR surrogates

BioAgeAccel transitions were also related to gradients in glucose, HbA1c, and 6 IR surrogates (Figure S6). Compared with the stable non-accelerated aging group, the accelerated-to-non-accelerated, non-accelerated-to-accelerated, and stable accelerated groups displayed average elevations of 0.009, 0.099, and 0.141, respectively, across the eight glycemic traits. When using the stable accelerated aging group as the reference, the accelerated-to-non-accelerated group showed an average decrease of approximately 0.133 across these glycemic traits. In particular, HbA1c exhibited the greatest decline of 0.24 (0.15∼0.32), whereas eGDR — a reverse marker of IR — showed a smaller but still significant increase of 0.06 (0∼0.12). Totally, PhenoAgeAccel demonstrated similar but weaker association patterns.

### Association of BioAgeAccel burdens with incident T2D, all-cause mortality, glucose, HbA1c and 6 IR surrogates

#### Association of BioAgeAccel burdens with incident T2D

BioAgeAccel burdens were significantly linked to T2D incidence, as demonstrated by the Kaplan-Meier curves that revealed evident differences in risk across tertiles of all burden indicators except for PhenoAgeAccel slope (*P*=0.090) (Figure S7). For KDMAccel, a steeper slope (per SD increase) was related to a 18.5% (6.8∼31.5%) higher risk, while the relative cumulative change could also elevate risk (HR=1.14 [1.03∼1.26]) and the cumulative measure showed an even greater effect (HR=1.25 [1.03∼1.51]) (Figure 3A). The three PhenoAgeAccel burdens demonstrated very similar influences on T2D risk, with an average HR of 1.24. When categorizing BioAgeAccel burdens into tertiles, we observed obvious dose-response relations although not all these trends were statistically significant (e.g., cumulative KDMAccel and PhenoAgeAccel) (Table S4).

**Figure 3.**
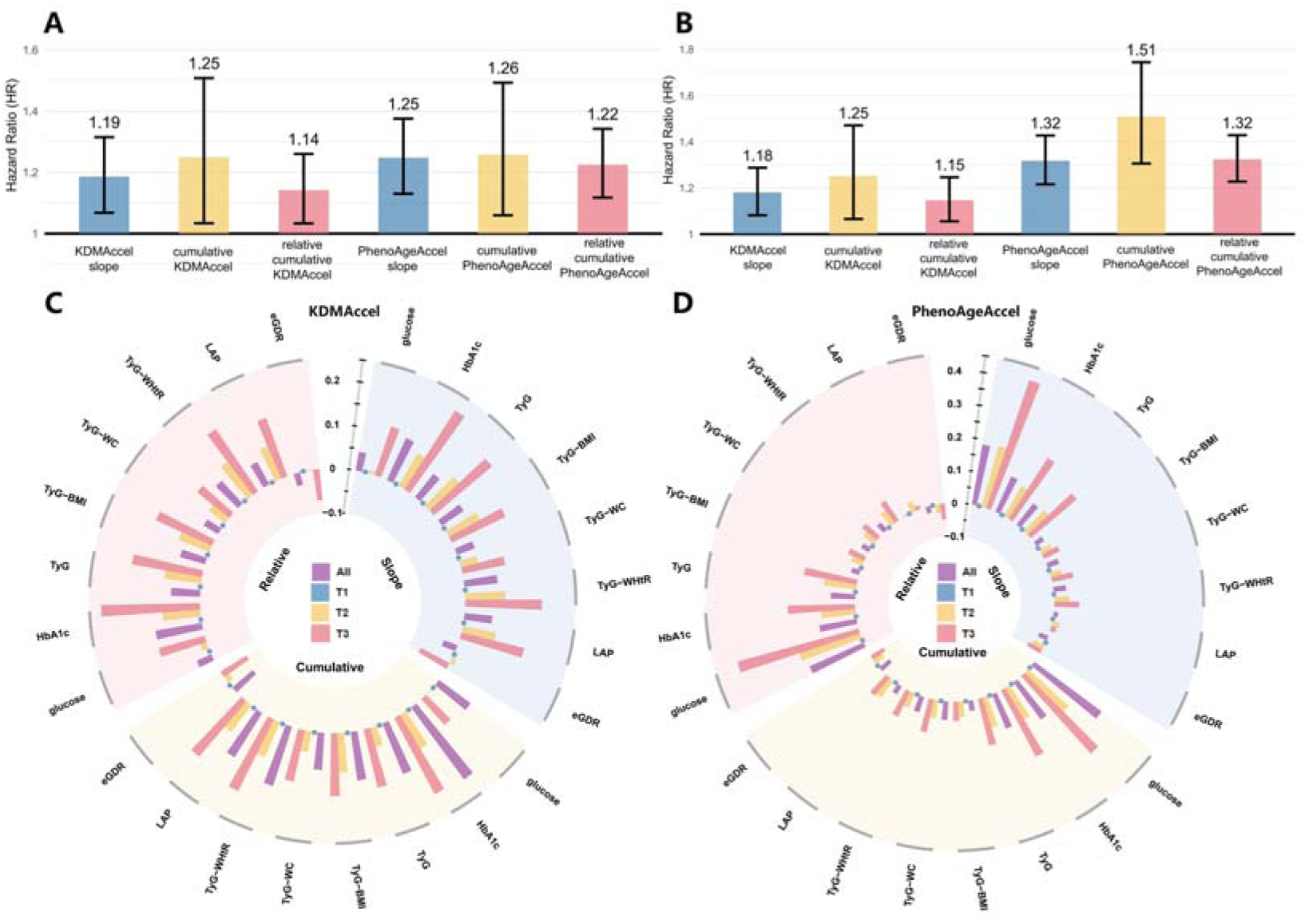
(A) Association of BioAgeAccel burdens with T2D; (B) Association of BioAgeAccel burdens with all-cause mortality; (C) Association of KDMAccel burdens with glycemic traits; (D) Association of PhenoAgeAccel burdens with glycemic traits. BioAgeAccel: biological aging acceleration; KDMAccel: Klemera-Doubal method biological age acceleration; PhenoAgeAccel: phenotypic age acceleration; T2D: type II diabetes; TyG: triglyceride-glucose; BMI: body mass index; WC: waist circumference; WHtR: waist-to-height ratio; LAP: lipid accumulation product; eGDR: estimated glucose disposal rate.

#### Association of BioAgeAccel burdens with all-cause mortality

BioAgeAccel burdens were also significantly associated with all-cause mortality, as illustrated by Kaplan-Meier curves showing clear differences in risk across tertiles of all burden indicators except for PhenoAgeAccel slope (*P*=0.878) (Figure S8). For KDMAccel, each SD increase in the slope, cumulative burden, and relative cumulative burden was associated with 18.0% (8.2∼28.7%), 25.2% (6.5∼47.1%), and 14.7% (5.5∼24.6%) higher risk of all-cause mortality (Figure 3B), respectively. Additionally, these PhenoAgeAccel burdens demonstrated very similar influences on mortality risk, with an average HR of 1.38 per SD increase. When categorizing BioAgeAccel burdens into tertiles, BioAgeAccel burdens also showed significant dose-response relationships with all-cause mortality risk (Table S5).

#### Association of BioAgeAccel burdens with incident glucose, HbA1c and 6 IR surrogates

BioAgeAccel burdens were also associated with the levels of glucose, HbA1c, and IR surrogates (Figure 3B-C), and the influences of BioAgeAccel burdens often presented a significant dose-response trend across tertiles. Among the KDMAccel burdens, HbA1c exhibited the most pronounced response, with the top tertile of cumulative burden associated with an increase of 0.20 (0.12∼0.27), and relative cumulative KDMAccel yielding an even greater increment of 0.22 (0.18∼0.27). Meanwhile, eGDR decreased significantly in the highest tertile of relative cumulative KDMAccel (-0.07 [-0.11∼-0.04]), consistent with elevated IR.

Consistent associations were also observed across PhenoAgeAccel burdens. The top tertile of PhenoAgeAccel slope was associated with a 0.41 (0.36∼0.46) increase in glucose, and a 0.22 (0.17∼0.27) rise in HbA1c. Relative cumulative PhenoAgeAccel demonstrated similar but slightly attenuated effects on these traits, while associations with LAP were generally weaker.

#### Robustness checks for incident T2D risk and all-cause mortality

Sensitivity analyses further supported the robustness of our findings described above. First, the exclusion of individuals occurring T2D within the first two years of follow-up yielded similar results, with both KDMAccel and PhenoAgeAccel retaining significant associations with incident T2D (Tables S6-S7). Similarly, sensitivity analyses for all-cause mortality produced consistent results after excluding early deaths, with BioAgeAccel transitions and burdens remaining significantly related to increased all-cause mortality risk (Tables S8-S9).

Second, removing participants receiving insulin or metformin therapies between baseline and the first follow-up did not substantially alter the associations (Tables S10-S11), though a slight loss of statistical significance was observed likely due to reduced power.

Third, to account for potential competing risk bias, we performed cause-specific hazard models by excluding individuals with non-T2D deaths. The results showed that, compared with individuals with stable non-accelerated aging (Table S12), those with stable accelerated aging displayed a significantly higher T2D risk (KDM: HR = 2.58 [2.00∼3.32]; PhenoAge: HR = 2.04 [1.38∼3.02]), while those transitioning from accelerated to non-accelerated aging still exhibited elevated risks, though to a lesser extent. Similar dose-response patterns were observed across burden indicators (Table S13).

#### Mediating roles of glycemic traits in the relation between BioAgeAccel and T2D

Our mediation analysis revealed that the eight glycemic traits played substantial but heterogeneous roles in mediating the effect of BioAgeAccel burden on T2D (Figure 4). We here ignored some glycemic traits (e.g., LAP) if the direction of the mediating effect was not consistent with that of the total effect. On average, six significant glycemic traits (glucose, HbA1c, TyG-BMI, TyG-WC, TyG-WHtR and eGDR) mediated 31.8% of the effect of KDMAccel slope and five significant glycemic traits (glucose, HbA1c, TyG-BMI, TyG-WC, TyG-WHtR) mediated 16.4% of the impact of PhenoAgeAccel slope. For cumulative burden, the mean mediation proportion of five significant glycemic traits (glucose, HbA1c, TyG-BMI, TyG-WHtR and eGDR) was 19.5% for KDMAccel and 23.8% for PhenoAgeAccel which also had five significant glycemic traits (glucose, HbA1c, TyG-WC, TyG-WHtR and eGDR) with mediating roles. For relative cumulative burden, the average mediation proportion rose to 24.4% and 17.6%, respectively, across six (except for TyG and LAP) or five (except for TyG, LAP and eGDR) significant glycemic traits. Totally, HbA1c and glucose exhibited the most pronounced mediating role in the relation between BioAgeAccel burden and T2D cross all these glycemic traits.

**Figure 4.**
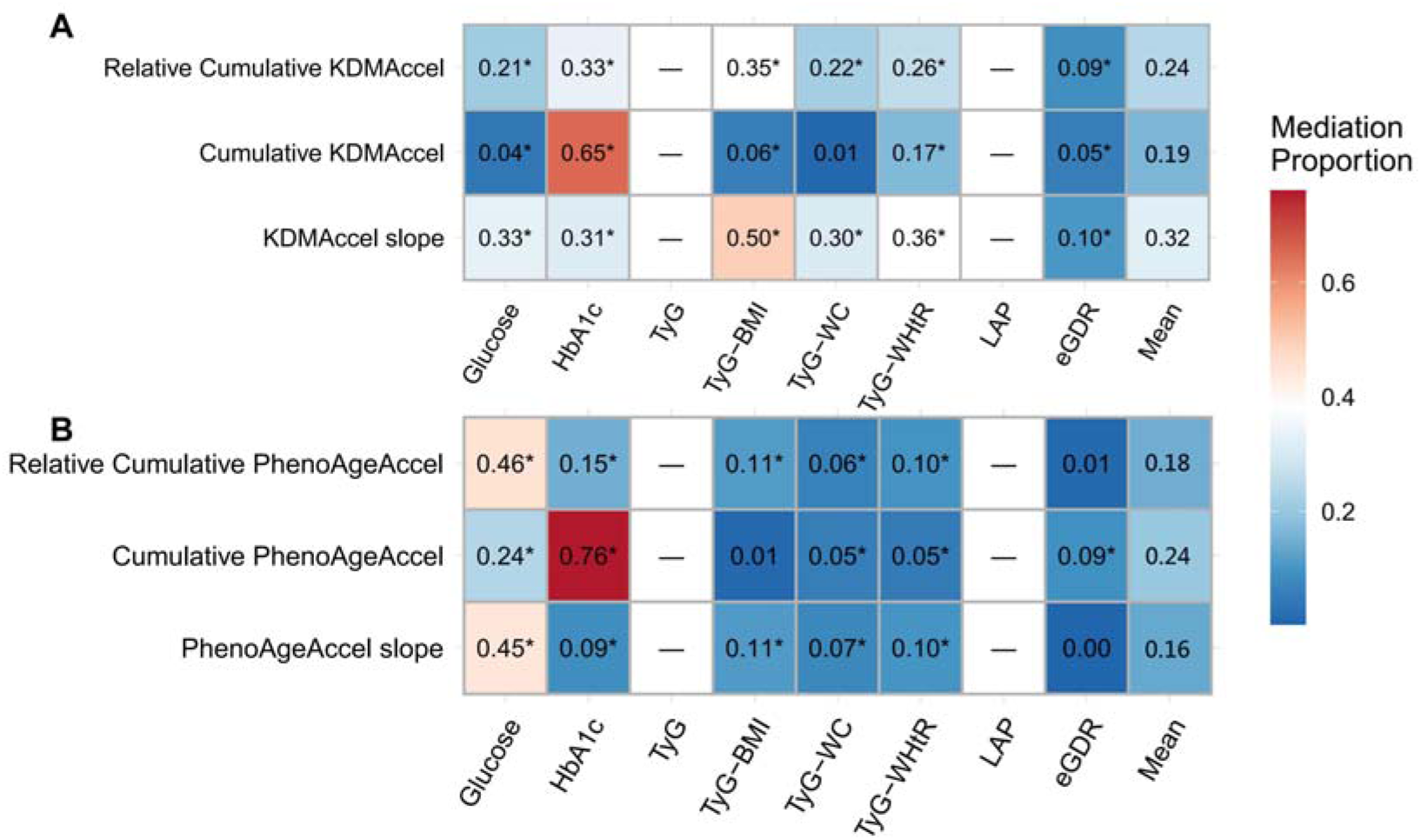
Mediation effects of glycemic traits on the association between BioAgeAccel and T2D. (A) KDMAccel burdens. (B) PhenoAgeAccel burdens. BioAgeAccel: biological aging acceleration; KDMAccel: Klemera-Doubal method biological age acceleration; PhenoAgeAccel: phenotypic age acceleration. T2D: type II diabetes. Heatmaps display the effect proportion mediated by each glycemic trait, representing the percentage of the total effect explained through the mediator pathway, with “*” indicating statistically significant mediation (joint significance test *P*<0.05), while “-” indicating that the direction of the mediating effect was not consistent with that of the total effect. The “Mean” column represents the average proportion mediated across all glycemic traits. All models were adjusted for age, sex, BMI, smoking status, drinking status, healthy diet score, physical activity, TDI, income, education, family history of T2D.

#### Predictive capacity of BioAgeAccel and FINDRISC on T2D

BioAgeAccel burdens and BioAgeAccel showed the potential to improve the predictive capacity of FINDRISC. For KDMAccel (Figure 5A), the C-index of FINDRISC achieved 0.712 (*se*=0.037), and increased to 0.738 (*se*=0.037) when including KDMAccel, representing a 2.6% relative improvement. While the inclusion of individual KDMAccel burdens (slope, cumulative KDMAccel, and relative cumulative KDMAccel) yielded comparable or even better enhancements, improving the C-index to 0.721 (*se*=0.037), 0.738 (*se*=0.037), and 0.720 (*se*=0.037), respectively. The best performance was observed for the model with FINDRISC, KDMAccel, and all KDMAccel burdens, achieving a C-index of 0.748 (*se*=0.036) — a 3.6% relative increase over FINDRISC and a 1.0% increase beyond using KDMAccel alone, underscoring the extra value of longitudinal aging burdens.

**Figure 5.**
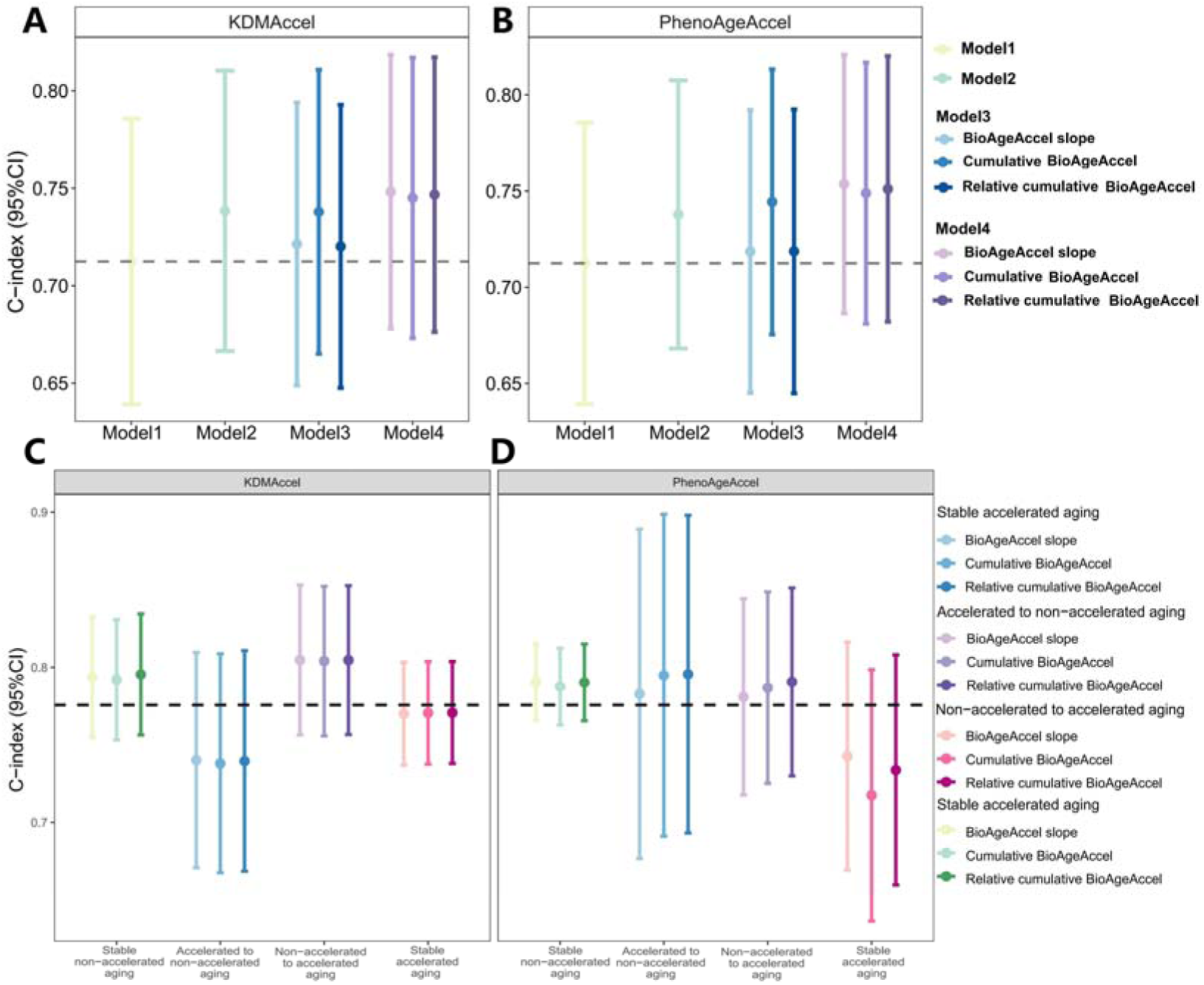
(A) Predictive capacity of KDMAccel, KDMAccel burdens and FINDSCORE on T2D; Model 1: with FINDRISC; Model 2: with FINDRISC and BioAgeAccel; Model 3: with FINDRISC and three BioAgeAccel burdens (slope, cumulative BioAgeAccel, and relative cumulative BioAgeAccel); Model 4: with FINDRISC, BioAgeAccel and three BioAgeAccel burdens. (B) Predictive capacity of PhenoAgeAccel, PhenoAgeAccel burdens and FINDSCORE on T2D; (C) Predictive capacity of Model 4 in KDMAccel transition statuses; (D) Predictive capacity of Model 4 in PhenoAgeAccel transition statuses. BioAgeAccel: biological aging acceleration; KDMAccel: Klemera-Doubal method biological age acceleration; PhenoAgeAccel: phenotypic age acceleration; FINDRISC: Finnish Diabetes Risk Score; T2D: type II diabetes.

For PhenoAgeAccel (Figure 5B), similar improvements were observed. The C-index rose from 0.712 for FINDRISC to 0.738 (*se*=0.036) (up by 2.6%) after incorporating PhenoAgeAccel. Importantly, cumulative PhenoAgeAccel showed an even greater predictive power, elevating the C-index to 0.744 (*se*=0.035) (3.2% gain). Among PhenoAgeAccel burdens, the slope and relative cumulative also contributed substantially (C-index=0.719 [*se*=0.038] and 0.719 [*se*=0.038], respectively). Again, the model including FINDRISC, PhenoAgeAccel, and all three burdens, achieved the highest C-index of 0.754 (*se*=0.034) — a 4.2% relative increase over FINDRISC, and a 1.6% increase beyond using PhenoAgeAccel alone.

Finally, using the optimal model incorporating FINDRISC, BioAgeAccel, and three BioAgeAccel burdens, we run the prediction model for each aging transition status. The best performance of KDMAccel was observed in the non-accelerated to accelerated aging group, with the C-index=0.805 (*se*=0.025) — standing for a 8.9% improvement compared with the accelerated to non-accelerated aging group (Figure 5C); whereas the best accuracy of PhenoAgeAccel was seen in the accelerated to non-accelerated aging group, with the C-index=0.796 (*se*=0.052) — representing a 10.9% improvement compared with the stable accelerated aging group (Figure 5D). These observations reflect prediction heterogeneity in aging statuses and highlight distinct risk characteristics during dynamic transitions in BioAgeAccel.

## Discussion

### Summary of our work

In this work, we have comprehensively evaluated the associations between BioAgeAccel, quantified by KDM and PhenoAge, and the risk of T2D, all-cause mortality and related glycemic traits. Individuals with accelerated biological aging, especially those maintaining or translating into acceleration over time, exhibited higher T2D and all-cause mortality risk, elevated glucose and HbA1c levels, and greater degrees of insulin resistance. Moreover, BioAgeAccel burdens were robustly linked to the future risk of T2D, all-cause mortality and also correlated with adverse glycemic profiles and insulin resistance indicators. We also revealed that some certain glycemic traits significantly mediated the influences of both BioAgeAccel itself and burdens. Finally, integrating BioAgeAccel into traditional diabetes risk score showed the ability to enhance predictive accuracy, especially in some specific aging transition statuses, implying its potential utility in early identification of individuals at high metabolic risk. The robustness of these findings was supported by several sensitivity analyses, long-term follow-up, large sample size, two complementary BioAgeAccel measures as well as comprehensive covariate adjustment.

### Comparison to previous studies

#### Dynamic biological aging trajectories and risks of T2D and all-cause mortality

Previous studies demonstrated that BioAgeAccel is a critical risk factor for T2D (Jiang, et al., 2024; Pan, et al., 2025; Yang, et al., 2025; Zhang, et al., 2025; Zhao and Yue, 2024), cardiometabolic multimorbidity, cardio-renal-metabolic multimorbidity and all-cause mortality (Fan, et al., 2025; He, et al., 2024; Jiang, et al., 2024; Tian, et al., 2025); however, all of them focused on static baseline measures, our work extended this paradigm by discovering that dynamic aging trajectories — not just static baseline acceleration — can also drive metabolic risk.

Recent experimental evidence has confirmed biological aging is a dynamic process that can be altered (Chen, et al., 2024; Poganik, et al., 2023). Our analysis revealed that BioAgeAccel transitions and cumulative burdens were significantly associated with T2D and all-cause mortality risk. Notably, we discovered participants progressing into or keeping accelerated aging exhibited substantially higher T2D and all-cause mortality risks compared with those with stable non-accelerated aging, while partial reversal (i.e., changing from accelerated aging to non-accelerated aging) likely alleviated but did not fully eliminate such risk. These findings imply the benefits of attenuating biological aging while demonstrating the lasting metabolic consequences of prior acceleration (Gladyshev, et al., 2021), highlighting the clinical significance of interventions targeting aging reversal.

Additionally, steeper slopes, greater cumulative burdens and relative cumulative increases in BioAgeAccel were dose-dependently related to T2D and all-cause mortality risk, emphasizing the importance of longitudinal aging trajectories relative to single timepoint for metabolic risk assessment. Our analysis also indicate that greater BioAgeAccel burdens are not only connected with increased risk of T2D and all-cause mortality but also to worse glycemic traits, further underscoring the role of BioAgeAccel in the development of T2D and related glycemic dysregulation.

Cause-specific hazard models accounting for competing risks confirmed that the association between BioAgeAccel (burden) and T2D remained significant even after excluding non-T2D deaths. These complementary results highlight the broader implications of biological aging on healthspan and lifespan, reinforcing its potential as a critical target for metabolic disease prevention.

In parallel, prior studies also suggested a potential bidirectional relationship, where T2D accelerated biological aging (Bahour, et al., 2022; Chiu, et al., 2024). Actually, T2D patients were often shown to have markedly higher biological age compared with non-diabetic controls, likely due to mechanisms involving chronic inflammation, oxidative stress, and glycemic dysregulation. These results demonstrate the complex interplay between biological aging and metabolic health, reinforcing the clinical value of monitoring dynamic aging processes to inform early interventions against T2D development.

#### Glycemic traits as mediators and early indicators along the aging-T2D

Our mediation analysis underscored the pivotal role of glycemic traits — particularly glucose and HbA1c — in bridging the association between BioAgeAccel and T2D, aligning with existing research that showed the importance of glycemic dysregulation in the pathogenesis of T2D (Miao, et al., 2024; Zimmet, et al., 2001). Our findings that these glycemic traits significantly mediated the relation between BioAgeAccel burden and T2D further reinforce their relevance. Notably, HbA1c emerged as the most potent mediator, consistent with its recognized status as a reliable indicator of long-term glycemic control and its predictive value for T2D risk (American Diabetes Association, 1989; American Diabetes Association, 2021).

The studied glycemic traits represent subclinical or prediabetic phenotypes (Abdul-Ghani and DeFronzo, 2009), and prior research has consistently shown their predictive value for T2D, even in the absence of diagnosed diabetes (Umeno, et al., 2018). Our results indicate that these glycemic traits not only serve as early indicators of subclinical metabolic deterioration but also as key intermediates in the progression from biological aging to overt diabetes, motivating our inclusion of them as secondary endpoints in analyses of BioAgeAccel transition and burden. Examining their associations with dynamic BioAgeAccel provides mechanistic and temporal insights into how biological aging manifests metabolically, supporting the biological mechanism that age-related metabolic deterioration may precede and contribute to the development of overt T2D (Tabák, et al., 2012).

#### Predictive performance differences

A recent study demonstrated that incorporating PhenoAgeAccel into risk models could improve the predictive performance of T2D risk, with the C-index rising from 0.662 to 0.679 (2.6% increase) (Pan, et al., 2025). Our work further advances this understanding by additionally incorporating dynamic measures of BioAgeAccel into the established FINDRISC model, with a significantly improved predictive accuracy of 3.6% for KDMAccel and 4.2% for PhenoAgeAccel, representing an approximately one-fold more enhancement compared with that work (Pan, et al., 2025). Further, we discovered that the predictive performance after integrating BioAgeAccel and its burdens into traditional diabetes risk score was distinct across various aging transition statuses, with more accuracy elevations observed in some specific aging transition statuses. Thus, our results reveal the added but differential predictive value of dynamic aging burdens in identifying individuals at high risk of T2D for various transition statuses, offering more precise directions of intervention and resource allocation.

### Potential clinical mechanisms

BioAgeAccel likely promotes T2D through interrelated pathways centered on metabolic and cellular dysfunction. Age-related adipose tissue senescence drives systemic insulin resistance via pro-inflammatory cytokine secretion (e.g., IL-6, TNF-α), which simultaneously stresses pancreatic β-cells and disrupts lipid homeostasis (Zhao and Yue, 2024). Concurrent mitochondrial inefficiency — marked by reduced ATP synthesis and elevated oxidative stress — compromises insulin-sensitive tissues while damaging β-cells, creating a self-reinforcing cycle of glucose dysregulation (Wiley and Campisi, 2016). These processes are exacerbated by chronic inflammation from senescence-associated secretory phenotypes (SASP), which persist even after transient BioAgeAccel reversal (Levine, et al., 2018), potentially contributing to residual T2D risk observed among recovered individuals who experienced the transition from accelerated to non-accelerated aging. Emerging evidence also implicates epigenetic dysregulation in aging-related metabolic decline, potentially altering insulin signaling and glucose transporter expression (Belsky, et al., 2015; Hillary, et al., 2020). Together, these mechanisms illustrate how BioAgeAccel disrupts metabolic equilibrium, with inflammatory, oxidative, and epigenetic pathways amplifying diabetes risk over time.

### Public health implications and advantages of this study

Our study offers several important clinical insights and methodological strengths that advance the understanding of biological aging in T2D prevention and risk stratification. First, our findings underscore the potential of biological aging markers, especially dynamic measures of BioAgeAccel, as early-warning indicators for T2D development. Unlike conventional glycemic traits such as glucose, HbA1c, or IR, BioAgeAccel offers a physiology-based, non-clinical indicator that can uncover latent or subclinical risk of T2D. It thus provides an alternative way to identify individuals at high risk of T2D, especially in some certain contexts such as asymptomatic or lean diabetes (Adam and Josten, 2008; George, et al., 2015), where glycemic traits may remain within normal ranges in early or atypical cases of diabetes, enabling earlier interventions targeting reversible aging pathways (Félix, et al., 2024). Crucially, even partial BioAgeAccel reversal reduced T2D risk, indicating modifiable aging as a preventive lever through lifestyle, pharmacotherapy, or social interventions.

Second, incorporating continuous glycemic traits such as glucose, HbA1c, and IR surrogates allows us to detect early impairments preceding overt T2D, thus enhancing our understanding of the connection between aging and diabetes. The integration of transition-based and burden-based BioAgeAccel metrics also provides novel insights into cumulative aging effects and significantly improved T2D risk prediction when added to FINDRISC. Moreover, we extended the clinical relevance of BioAgeAccel beyond T2D by demonstrating its significant associations with all-cause mortality. This finding aligns with growing evidence that accelerated biological aging not only increases chronic disease risk but also shortens lifespan.

### Limitations of this work

Several limitations should be acknowledged. First, although our study benefits from repeated measures of markers, the follow-up interval remains limited to two timepoints, which may not fully capture long-term fluctuations. Second, the calculation of BioAge relied on selected clinical markers, which, although widely used, may not reflect all dimensions of biological aging such as molecular or cellular-level changes (Rutledge, et al., 2022). Third, the sample sizes of incident cases and all-cause deaths were relatively insufficient, reducing statistical power to detect associations with dynamic changes in BioAge. Lastly, the first follow-up samples were drawn from volunteers willing to participate in intensive imaging assessments, potentially introducing a healthy volunteer bias — these participants were generally healthier, of higher socioeconomic status, and more likely to exhibit favorable health behaviors compared with the full baseline cohort (Fry, et al., 2017). This selective participation may limit the generalizability of our longitudinal findings to broader populations.

### Conclusions

Our study demonstrated that BioAgeAccel, assessed by KDM and PhenoAge, and its burdens were significantly related to increased risk of T2D and all-cause mortality as well as worse glycemic and insulin resistance profiles. Incorporating BioAgeAccel and the burdens into T2D risk models improved prediction beyond traditional tools.

## Ethics approval and informed consent

The UK Biobank had approval from the North West Multi-Centre Research Ethics Committee (MREC) as a Research Tissue Bank (RTB) approval. All participants provided written informed consent before enrolment in the study, which was conducted in accordance with the Declaration of Helsinki. This approval means that researchers do not require separate ethical clearance and can operate under the RTB approval.

## Consent for publication

All authors have approved the manuscript and given their consent for submission and publication.

## Data availability

All data generated or analyzed during this study are included in this published article and its supplementary information files. This study used the UK Biobank resource with the application ID 88159. Researchers can access to the UK Biobank by submitting an application to the UK Biobank official website (https://www.ukbiobank.ac.uk/).

## Author’s contributions

PZ conceived the idea for the study. PZ obtained the data. PZ and YY, CZ cleared up the datasets; YY performed the data analyses. PZ, YY, and CZ interpreted the results of the data analyses. PZ, YY, and CZ wrote the manuscript with the help from other authors.

## Acknowledgements

This study was mainly based on the UK Biobank resource under application number 88159. The UK Biobank was established by the Wellcome Trust medical charity, Medical Research Council, Department of Health, Scottish Government, and the Northwest Regional Development Agency. It has also had funding from the Welsh Assembly Government, British Heart Foundation and Diabetes UK. The data analyses in the present study were carried out with the high-performance computing cluster that was supported by the special central finance project of local universities for Xuzhou Medical University.

## References

Abdul-Ghani, M.A. and DeFronzo, R.A. (2009) Pathophysiology of prediabetes, Current diabetes reports, 9, 193–199.

Adam, J.M. and Josten, D. (2008) Isolated post-challenge hyperglycemia: concept and clinical significance, Acta medica Indonesiana, 40, 171–175.

Ahadi, S., et al. (2020) Personal aging markers and ageotypes revealed by deep longitudinal profiling, Nature medicine, 26, 83–90.

American Diabetes Association (1989) Standards of medical care for patients with diabetes mellitus, Diabetes care, 12, 365–368.

American Diabetes Association (2021) 2. Classification and Diagnosis of Diabetes: Standards of Medical Care in Diabetes-2021, Diabetes care, 44, S15-s33.

Andersen, P.K., et al. (2012) Competing risks in epidemiology: possibilities and pitfalls, International journal of epidemiology, 41, 861–870.

Bahour, N., et al. (2022) Diabetes mellitus correlates with increased biological age as indicated by clinical biomarkers, GeroScience, 44, 415–427.

Belsky, D.W., et al. (2015) Quantification of biological aging in young adults, Proceedings of the National Academy of Sciences of the United States of America, 112, E4104–4110.

Boonpor, J., et al. (2023) Dose-response relationship between device-measured physical activity and incident type 2 diabetes: findings from the UK Biobank prospective cohort study, BMC Med, 21, 191.

Brownlee, M. (2005) The pathobiology of diabetic complications: a unifying mechanism, Diabetes, 54, 1615–1625.

Chen, X., et al. (2024) Small extracellular vesicles from young plasma reverse age-related functional declines by improving mitochondrial energy metabolism, Nature aging, 4, 814–838.

Chiu, D.T., et al. (2024) Essential Nutrients, Added Sugar Intake, and Epigenetic Age in Midlife Black and White Women: NIMHD Social Epigenomics Program, JAMA network open, 7, e2422749.

Cui, H., et al. (2022) Cumulative triglyceride-glucose index is a risk for CVD: a prospective cohort study, Cardiovasc. Diabetol., 21, 22.

DeFronzo, R.A., et al. (2015) Type 2 diabetes mellitus, Nature reviews. Disease primers, 1, 15019.

Er, L.K., et al. (2016) Triglyceride Glucose-Body Mass Index Is a Simple and Clinically Useful Surrogate Marker for Insulin Resistance in Nondiabetic Individuals, PloS one, 11, e0149731.

Félix, J., et al. (2024) Frailty and biological age. Which best describes our aging and longevity?, Molecular aspects of medicine, 98, 101291.

Fan, G., et al. (2025) Reproductive factors and biological aging: the association with all-cause and cause-specific premature mortality, Human reproduction (Oxford, England), 40, 148–156.

Fry, A., et al. (2017) Comparison of Sociodemographic and Health-Related Characteristics of UK Biobank Participants With Those of the General Population, American journal of epidemiology, 186, 1026–1034.

George, A.M., Jacob, A.G. and Fogelfeld, L. (2015) Lean diabetes mellitus: An emerging entity in the era of obesity, World journal of diabetes, 6, 613–620.

Gladyshev, V.N., et al. (2021) Molecular Damage in Aging, Nature aging, 1, 1096–1106.

He, D., et al. (2024) Changes in frailty and incident cardiovascular disease in three prospective cohorts, Eur. Heart J., 45, 1058–1068.

He, Y., et al. (2024) Accelerated biological aging: unveiling the path to cardiometabolic multimorbidity, dementia, and mortality, Frontiers in public health, 12, 1423016.

Hillary, R.F., et al. (2020) Epigenetic measures of ageing predict the prevalence and incidence of leading causes of death and disease burden, Clinical epigenetics, 12, 115.

International Expert Committee (2009) International Expert Committee report on the role of the A1C assay in the diagnosis of diabetes, Diabetes care, 32, 1327–1334.

Jiang, M., et al. (2024) Accelerated biological aging elevates the risk of cardiometabolic multimorbidity and mortality, Nature cardiovascular research, 3, 332–342.

Jylhävä, J., Pedersen, N.L. and Hägg, S. (2017) Biological Age Predictors, EBioMedicine, 21, 29–36.

Kahn, H.S. (2005) The “lipid accumulation product” performs better than the body mass index for recognizing cardiovascular risk: a population-based comparison, BMC cardiovascular disorders, 5, 26.

Klemera, P. and Doubal, S. (2006) A new approach to the concept and computation of biological age, Mechanisms of ageing and development, 127, 240–248.

Kolb, H. and Martin, S. (2017) Environmental/lifestyle factors in the pathogenesis and prevention of type 2 diabetes, BMC medicine, 15, 131.

Kuo, C.L., et al. (2021) Genetic associations for two biological age measures point to distinct aging phenotypes, Aging cell, 20, e13376.

Levine, M.E. (2013) Modeling the rate of senescence: can estimated biological age predict mortality more accurately than chronological age?, The journals of gerontology. Series A, Biological sciences and medical sciences, 68, 667–674.

Levine, M.E., et al. (2018) An epigenetic biomarker of aging for lifespan and healthspan, Aging, 10, 573–591.

Li, X., et al. (2021) Obesity and the relation between joint exposure to ambient air pollutants and incident type 2 diabetes: A cohort study in UK Biobank, PLoS medicine, 18, e1003767.

Lim, J., et al. (2019) Comparison of triglyceride glucose index, and related parameters to predict insulin resistance in Korean adults: An analysis of the 2007-2010 Korean National Health and Nutrition Examination Survey, PloS one, 14, e0212963.

Lindström, J. and Tuomilehto, J. (2003) The diabetes risk score: a practical tool to predict type 2 diabetes risk, Diabetes care, 26, 725–731.

Liu, Z., et al. (2018) A new aging measure captures morbidity and mortality risk across diverse subpopulations from NHANES IV: A cohort study, PLoS medicine, 15, e1002718.

Miao, K., et al. (2024) Association between epigenetic age and type 2 diabetes mellitus or glycemic traits: A longitudinal twin study, Aging cell, 23, e14175.

Office for National Statistics (2017) Median Gross Income, All Households 1977 to Financial Year Ending 2016.

Ong, K.L., et al. (2023) Global, regional, and national burden of diabetes from 1990 to 2021, with projections of prevalence to 2050: a systematic analysis for the Global Burden of Disease Study 2021, The Lancet, 402, 203–234.

Pan, L., et al. (2025) Association of accelerated phenotypic aging, genetic risk, and lifestyle with progression of type 2 diabetes: a prospective study using multi-state model, BMC medicine, 23, 62.

Peng, Y., et al. (2023) Association between the Finnish Diabetes Risk Score and cancer in middle-aged and older adults: Involvement of inflammation, Metabolism: clinical and experimental, 144, 155586.

Poganik, J.R., et al. (2023) Biological age is increased by stress and restored upon recovery, Cell metabolism, 35, 807–820.e805.

Ramdas Nayak, V.K., et al. (2022) Triglyceride Glucose (TyG) Index: A surrogate biomarker of insulin resistance, *JPMA*. The Journal of the Pakistan Medical Association, 72, 986–988.

Rutledge, J., Oh, H. and Wyss-Coray, T. (2022) Measuring biological age using omics data, Nature reviews. Genetics, 23, 715–727.

Shi, B., et al. (2021) CMAverse: A Suite of Functions for Reproducible Causal Mediation Analyses, Epidemiology (Cambridge, Mass.), 32, e20–e22.

Sudlow, C., et al. (2015) UK biobank: an open access resource for identifying the causes of a wide range of complex diseases of middle and old age, PLoS Med, 12, e1001779.

Sun, Y., et al. (2024) Long-term changes in frailty and incident type 2 diabetes: A prospective cohort study based on the UK Biobank, Diabetes Obes. Metab., 26, 3352–3360.

Suzuki, K., et al. (2024) Genetic drivers of heterogeneity in type 2 diabetes pathophysiology, Nature, 627, 347–357.

Tabák, A.G., et al. (2012) Prediabetes: a high-risk state for diabetes development, Lancet (London, England), 379, 2279–2290.

Tian, Y., et al. (2025) Biological Age Acceleration Associated with the Progression Trajectory of Cardio-Renal-Metabolic Multimorbidity: A Prospective Cohort Study, Nutrients, 17.

Umeno, A., et al. (2018) Early diagnosis of type 2 diabetes based on multiple biomarkers and non-invasive indices, Journal of clinical biochemistry and nutrition, 62, 187–194.

Valeri, L. and VanderWeele, T.J. (2013) Mediation Analysis Allowing for Exposure-Mediator Interactions and Causal Interpretation: Theoretical Assumptions and Implementation With SAS and SPSS Macros, Psychological Methods, 18, 137–150.

van Buuren, S. and Groothuis-Oudshoorn, K. (2011) mice: Multivariate Imputation by Chained Equations in R, J. Stat. Softw., 45, 1–67.

VanderWeele, T.J. and Vansteelandt, S. (2014) Mediation Analysis with Multiple Mediators, Epidemiologic methods, 2, 95–115.

Wang, M., et al. (2021) Joint exposure to various ambient air pollutants and incident heart failure: a prospective analysis in UK Biobank, Eur. Heart J., 42, 1582–1591.

Wiley, C.D. and Campisi, J. (2016) From Ancient Pathways to Aging Cells-Connecting Metabolism and Cellular Senescence, Cell metabolism, 23, 1013–1021.

Xiang, H., et al. (2024) Clinical biomarker-based biological ageing and the risk of adverse outcomes in patients with chronic kidney disease, Age and ageing, 53.

Xiang, Y., et al. (2024) Tea consumption and attenuation of biological aging: a longitudinal analysis from two cohort studies, Lancet Reg Health West Pac, 42, 100955.

Yang, G., et al. (2025) Biological age acceleration and interaction with genetic predisposition in the risk of type 2 diabetes and coronary artery disease, GeroScience.

Zeng, P., Shao, Z. and Zhou, X. (2021) Statistical methods for mediation analysis in the era of high-throughput genomics: Current successes and future challenges, Computational and structural biotechnology journal, 19, 3209–3224.

Zhang, Y., et al. (2021) Association Between Cumulative Low-Density Lipoprotein Cholesterol Exposure During Young Adulthood and Middle Age and Risk of Cardiovascular Events, JAMA Cardiology, 6, 1406–1413.

Zhang, Z., et al. (2025) Accelerated biological aging, mediating amino acids, and risk of incident type 2 diabetes: a prospective cohort study, Journal of endocrinological investigation, 48, 435–443.

Zhang, Z., et al. (2024) Insulin resistance assessed by estimated glucose disposal rate and risk of incident cardiovascular diseases among individuals without diabetes: findings from a nationwide, population based, prospective cohort study, Cardiovasc. Diabetol., 23, 194.

Zhao, Y. and Yue, R. (2024) Aging adipose tissue, insulin resistance, and type 2 diabetes, Biogerontology, 25, 53–69.

Zheng, S., et al. (2016) Triglyceride glucose-waist circumference, a novel and effective predictor of diabetes in first-degree relatives of type 2 diabetes patients: cross-sectional and prospective cohort study, Journal of translational medicine, 14, 260.

Zimmet, P., Alberti, K.G. and Shaw, J. (2001) Global and societal implications of the diabetes epidemic, Nature, 414, 782–787.

