## Supplementary File for "Longitudinal insights into dynamic patterns and cumulative burdens of biological age acceleration in relation to type II diabetes mellitus, all-cause mortality and glycemic traits"

**Assessment of IR surrogates**

To comprehensively assess insulin resistance, we derived several validated insulin resistance (IR) surrogate indices based on routinely measured biomarkers and anthropometric variables available in the UK Biobank. All biomarkers were collected during the second follow-up visit to ensure data completeness and temporal proximity.

The following IR surrogates, including triglyceride-glucose (TyG) (Ramdas Nayak, et al., 2022), TyG combined with body mass index (BMI) (TyG-BMI) (Er, et al., 2016), TyG combined with waist circumference (WC) (TyG-WC) (Zheng, et al., 2016), TyG combined with waist-to-height ratio (WHtR) (TyG-WHtR) (Lim, et al., 2019), lipid accumulation product (LAP), which reflects lipid overaccumulation and is used as a marker of IR in epidemiologic studies (Kahn, 2005), and the estimated glucose disposal rate (eGDR) (Zhang, et al., 2024), were calculated:

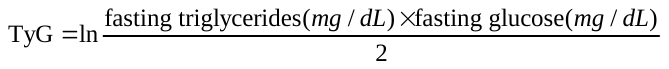

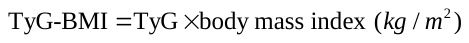

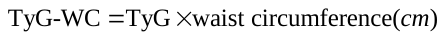

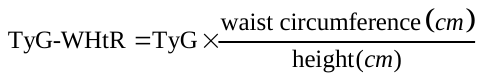

For men:
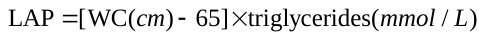

For women:
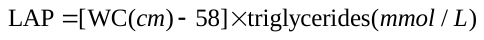

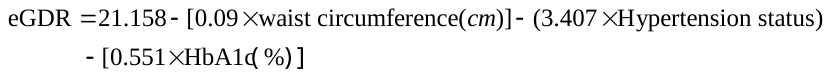

Participants with complete data on the above biomarkers and anthropometric measurements at the second follow-up were included in the IR surrogate analyses.

**Assessment of accelerated biological aging**

The Klemera-Doubal method biological age (KDM) is a technique that estimates BA using a combination of chronological age (CA) and nine biomarkers: albumin, alkaline phosphatase, blood urea nitrogen, creatinine, C-reactive protein, forced expiratory volume in one second, glycated hemoglobin, systolic blood pressure, and total cholesterol (Kwon and Belsky, 2021). Model training and calibration were conducted using data from the National Health and Nutrition Examination Survey III (NHANES III) (Mak, et al., 2023). The formula for KDM can be expressed as

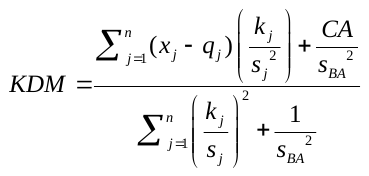

where *x_j_* represents the *j^th^* biomarker; *q*, *k*, and *s* represent the intercept, slope, and root mean squared error when regressed the *j^th^* biomarker on *CA,* respectively; *s_BA_* represents a scaling factor equal to the square root of the variance in chronological age explained by the biomarker set. To correct the skewed distribution of clinical biomarkers, we performed truncated extremes for each biomarker, setting the bottom 1% of biomarker values to the 1^st^ percentile, and the top 1% to the 99^th^ percentile. KDM-BA and KDM-BA acceleration were computed via the R BioAge package (van Buuren and Groothuis-Oudshoorn, 2011).

The PhenoAge algorithm is derived from multivariate analysis of mortality hazards. The original PhenoAge algorithm was constructed from elastic-net Gompertz regression of mortality on 42 biomarkers in the Third National Health and Nutrition Examination Survey (NHANES III) (Levine, et al., 2018). This analysis selected nine biomarkers: albumin, alkaline phosphatase, creatinine, C-reactive protein, glucose, mean cell volume, red cell distribution width, white blood cell count, and lymphocyte proportion, and chronological age (Liu, et al., 2018). The formula is

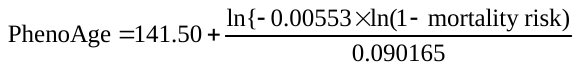

where

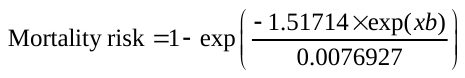

and

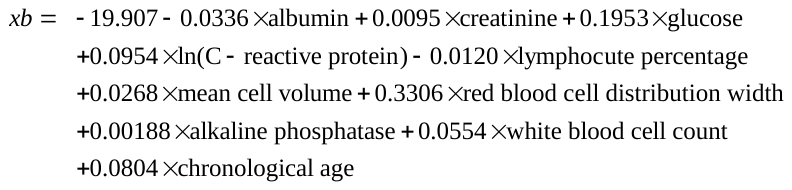

**Calculation of FINDRISC**

Finnish Diabetes Risk Score (FINDRISC) was calculated for each participant using self-reported and measured variables available in the UK Biobank, following the original FINDRISC algorithm developed by the Finnish Diabetes Association. This non-invasive score estimates the 10-year risk of developing type II diabetes mellitus (T2D). Specific coefficients of models and algorithms are presented in Table S14. The total FINDRISC score ranges from 0 to 26, with higher scores indicating a greater risk of developing T2D. In our study, we derived each component based on the closest available data in the UK Biobank baseline assessment and applied the standardized scoring algorithm to obtain an individual-level FINDRISC. Participants with complete data on all eight variables were included in the analysis.

Table S1. Definitions and sources of information for type II diabetes, glucose and HbA1c in the UK Biobank.

| Diseases | Fields | Codes |
| --- | --- | --- |
| T2D | 41270 | E11 |
|  | 20002 | 1223 |
|  | 2443 | 1 |
|  | 6153 | 3 |
|  | 6157 | 3 |
|  | 2976 | 3 |
| Glucose | 30740 | - |
| HbA1c | 30750 | - |

Table S2. Baseline characteristics of included participants in the UK Biobank study according to KDMAccel transitions.

| Characteristics | Stable non-accelerated  aging | Non-accelerated to  accelerated aging | Accelerated to  non-accelerated aging | Stable accelerated  aging |
| --- | --- | --- | --- | --- |
| **Number of participants (%)** | 6744 (49.0) | 952 (6.9) | 2925 (21.3) | 3130 (22.8) |
| **Baseline KDMAccel** | -2.8 (1.7) | 1.2 (1.1) | -1.6 (1.3) | 1.7 (1.4) |
| **Age** | 62.2 (7.4) | 62.6 (7.2) | 61.2 (7.4) | 61.1 (7.4) |
| **Body mass index** | 25.9 (3.7) | 27.5 (4.4) | 26.7 (4.2) | 28.2 (5.1) |
| **Townsend deprivation index** | -2.2 (2.6) | -1.9 (2.8) | -2 (2.7) | -1.8 (2.9) |
| **Systolic blood pressure** | 132.6 (16) | 146.6 (18.9) | 134.5 (15.9) | 147.3 (18.5) |
| **Diastolic blood pressure** | 79.5 (9.4) | 85.8 (10.1) | 80.5 (9.2) | 85.5 (9.6) |
| **Gender** | | | | |
| Female, *N* (%) | 2776 (41.2) | 442 (46.4) | 1795 (61.4) | 2036 (65.0) |
| Male, *N* (%) | 3968 (58.8) | 510 (53.6) | 1130 (38.6) | 1094 (35.0) |
| **Smoking status** | | | | |
| No, *N* (%) | 2486 (36.9) | 348 (36.6) | 1149 (39.3) | 1273 (40.7) |
| Yes, *N* (%) | 4255 (63.1) | 604 (63.4) | 1776 (60.7) | 1855 (59.3) |
| Missing, *N* (%) | 3 (0.0) | 0 (0.0) | 0 (0.0) | 2 (0.1) |
| **Alcohol consumption** | | | | |
| No, *N* (%) | 138 (2.0) | 27 (2.8) | 76 (2.6) | 101 (3.2) |
| Yes, *N* (%) | 6606 (98.0) | 925 (97.2) | 2849 (97.4) | 3029 (96.8) |
| **Healthy diet score** | | | | |
| 0, *N* (%) | 69 (1.0) | 11 (1.2) | 31 (1.1) | 42 (1.3) |
| 1, *N* (%) | 365 (5.4) | 66 (6.9) | 180 (6.2) | 249 (8) |
| 2, *N* (%) | 997 (14.8) | 168 (17.6) | 426 (14.6) | 525 (16.8) |
| 3, *N* (%) | 1829 (27.1) | 282 (29.6) | 768 (26.3) | 858 (27.4) |
| 4, *N* (%) | 2063 (30.6) | 259 (27.2) | 909 (31.1) | 926 (29.6) |
| 5, *N* (%) | 1223 (18.1) | 137 (14.4) | 517 (17.7) | 427 (13.6) |
| Missing, *N* (%) | 198 (2.9) | 29 (3.0) | 94 (3.2) | 103 (3.3) |
| **Physical activity** | | | | |
| Low, *N* (%) | 1063 (15.8) | 170 (17.9) | 454 (15.5) | 527 (16.8) |
| Moderate, N (%) | 2417 (35.8) | 320 (33.6) | 1004 (34.3) | 1050 (33.5) |
| High, *N* (%) | 2358 (35.0) | 302 (31.7) | 978 (33.4) | 945 (30.2) |
| Missing, *N* (%) | 906 (13.4) | 160 (16.8) | 489 (16.7) | 608 (19.4) |
| **Income** | | | | |
| <£18,000, *N* (%) | 607 (9.0) | 113 (11.9) | 317 (10.8) | 421 (13.5) |
| £18,000~£30,999, *N* (%) | 1602 (23.8) | 263 (27.6) | 737 (25.2) | 829 (26.5) |
| £31,000~£51,999, *N* (%) | 2005 (29.7) | 258 (27.1) | 807 (27.6) | 862 (27.5) |
| £52,000~£100,000, *N* (%) | 1767 (26.2) | 221 (23.2) | 756 (25.8) | 683 (21.8) |
| >£100,000, *N* (%) | 525 (7.8) | 57 (6.0) | 169 (5.8) | 158 (5.0) |
| Missing, *N* (%) | 238 (3.5) | 40 (4.2) | 139 (4.8) | 177 (5.7) |
| **Education level** | | | | |
| High school or below, *N* (%) | 3215 (47.7) | 546 (57.4) | 1605 (54.9) | 1832 (58.5) |
| College degree or above, *N* (%) | 3527 (52.3) | 404 (42.4) | 1320 (45.1) | 1295 (41.4) |
| Missing, *N* (%) | 2 (0.0) | 2 (0.2) | 0 (0.0) | 3 (0.1) |
| **Family history of T2D** | | | | |
| No, *N* (%) | 5168 (76.6) | 711 (74.7) | 2154 (73.6) | 2256 (72.1) |
| Yes, *N* (%) | 1576 (23.4) | 241 (25.3) | 771 (26.4) | 874 (27.9) |

Note: KDMAccel: Klemera-Doubal method biological age acceleration; T2D: type II diabetes. Continuous variables are described as mean (SD), and categorical variables are described as frequency (percentage).

Table S3. Baseline characteristics of included participants in the UK Biobank study according to PhenoAgeAccel transitions.

| Characteristics | Stable non-accelerated  aging | Non-accelerated to  accelerated aging | Accelerated to  non-accelerated aging | Stable accelerated  aging |
| --- | --- | --- | --- | --- |
| **Number of participants (%)** | 12052 (87.6) | 363 (2.6) | 1011 (7.4) | 325 (2.4) |
| **Baseline PhenoAgeAccel** | -7.5 (3.1) | 2.0 (1.9) | -4.2 (2.7) | 3.1 (2.8) |
| **Age** | 61.7 (7.3) | 61.1 (8.0) | 62.1 (7.9) | 62.4 (8.1) |
| **Body mass index** | 26.5 (4.1) | 28.1 (5.5) | 27.8 (5.1) | 28.6 (5.8) |
| **Townsend deprivation index** | -2.1 (2.6) | -1.8 (2.9) | -1.6 (3.0) | -1.3 (3.2) |
| **Systolic blood pressure** | 137.2 (18.0) | 137.9 (18.2) | 138.2 (17.8) | 138.7 (17.5) |
| **Diastolic blood pressure** | 81.4 (9.8) | 81.2 (10.1) | 82.4 (9.8) | 81.5 (9.4) |
| **Gender** | | | | |
| Female, *N* (%) | 6338 (52.6) | 186 (51.2) | 399 (39.5) | 126 (38.8) |
| Male, *N* (%) | 5714 (47.4) | 177 (48.8) | 612 (60.5) | 199 (61.2) |
| **Smoking status** | | | | |
| No, *N* (%) | 4658 (38.6) | 133 (36.6) | 355 (35.1) | 110 (33.8) |
| Yes, *N* (%) | 7390 (61.3) | 230 (63.4) | 656 (64.9) | 214 (65.8) |
| Missing, *N* (%) | 4 (0.1) | 0 (0.0) | 0 (0.0) | 1 (0.3) |
| **Alcohol consumption** | | | | |
| No, *N* (%) | 298 (2.5) | 10 (2.8) | 22 (2.2) | 12 (3.7) |
| Yes, *N* (%) | 11754 (97.5) | 353 (97.2) | 989 (97.8) | 313 (96.3) |
| **Healthy diet score** | | | | |
| 0, *N* (%) | 112 (0.9) | 8 (2.2) | 19 (1.9) | 14 (4.3) |
| 1, *N* (%) | 716 (5.9) | 37 (10.2) | 72 (7.1) | 35 (10.8) |
| 2, *N* (%) | 1779 (14.8) | 66 (18.2) | 196 (19.4) | 75 (23.1) |
| 3, *N* (%) | 3254 (27.0) | 93 (25.6) | 309 (30.6) | 81 (24.9) |
| 4, *N* (%) | 3713 (30.8) | 107 (29.5) | 262 (25.9) | 75 (23.1) |
| 5, *N* (%) | 2113 (17.5) | 39 (10.7) | 119 (11.8) | 33 (10.2) |
| Missing, *N* (%) | 365 (3.0) | 13 (3.6) | 34 (3.4) | 12 (3.7) |
| **Physical activity** | | | | |
| Low, *N* (%) | 1880 (15.6) | 78 (21.5) | 179 (17.7) | 77 (23.7) |
| Moderate, N (%) | 4208 (34.9) | 125 (34.4) | 355 (35.1) | 103 (31.7) |
| High, *N* (%) | 4097 (34.0) | 94 (25.9) | 307 (30.4) | 85 (26.2) |
| Missing, *N* (%) | 1867 (15.5) | 66 (18.2) | 170 (16.8) | 60 (18.5) |
| **Income** | | | | |
| <£18,000, *N* (%) | 1204 (10.0) | 38 (10.5) | 155 (15.3) | 61 (18.8) |
| £18,000~£30,999, *N* (%) | 2947 (24.5) | 106 (29.2) | 267 (26.4) | 111 (34.2) |
| £31,000~£51,999, *N* (%) | 3479 (28.9) | 102 (28.1) | 275 (27.2) | 76 (23.4) |
| £52,000~£100,000, *N* (%) | 3073 (25.5) | 86 (23.7) | 219 (21.7) | 49 (15.1) |
| >£100,000, *N* (%) | 834 (6.9) | 15 (4.1) | 51 (5.0) | 9 (2.8) |
| Missing, *N* (%) | 515 (4.3) | 16 (4.4) | 44 (4.4) | 19 (5.8) |
| **Education level** | | | | |
| High school or below, *N* (%) | 6156 (51.1) | 223 (61.4) | 605 (59.8) | 214 (65.8) |
| College degree or above, *N* (%) | 5890 (48.9) | 140 (38.6) | 405 (40.1) | 111 (34.2) |
| Missing, *N* (%) | 6 (0.0) | 0 (0.0) | 1 (0.1) | 0 (0.0) |
| **Family history of T2D** | | | | |
| No, *N* (%) | 9044 (75.0) | 261 (71.9) | 751 (74.3) | 233 (71.7) |
| Yes, *N* (%) | 3008 (25.0) | 102 (28.1) | 260 (25.7) | 92 (28.3) |

Note: PhenoAgeAccel: PhenoAge acceleration; T2D: type II diabetes. Continuous variables are described as mean (SD), and categorical variables are described as frequency (percentage).

Table S4. Association of BioAgeAccel burdens with the risk of incident T2D.

| BioAgeAccel burdens | | Events | HR (95%CI) | *P*-value |
| --- | --- | --- | --- | --- |
| KDMAccel slope | T1 | 158/4426 | 1.00 (Ref.) | - |
|  | T2 | 108/4475 | 0.93 (0.72~1.19) | 0.545 |
|  | T3 | 146/4438 | 1.48 (1.15~1.89) | 1.94×10^-4^ |
|  | *P* for trend | 2.84×10^-3^ | | |
| Cumulative KDMAccel | T1 | 79/4505 | 1.00 (Ref.) | - |
|  | T2 | 110/4473 | 1.01 (0.74~1.39) | 0.944 |
|  | T3 | 223/4361 | 1.32 (0.89~1.94) | 0.166 |
|  | *P* for trend | 0.123 | | |
| Relative cumulative KDMAccel | T1 | 156/4428 | 1.00 (Ref.) | - |
|  | T2 | 106/4477 | 0.94 (0.73~1.21) | 0.646 |
|  | T3 | 150/4434 | 1.46 (1.15~1.87) | 2.08×10^-3^ |
|  | *P* for trend | 2.61×10^-3^ | | |
| PhenoAgeAccel slope | T1 | 134/4450 | 1.00 (Ref.) | - |
|  | T2 | 118/4465 | 1.09 (0.85~1.41) | 0.491 |
|  | T3 | 160/4424 | 1.59 (1.24~2.04) | 2.81×10^-4^ |
|  | *P* for trend | 2.20×10^-4^ | | |
| Cumulative PhenoAgeAccel | T1 | 81/4503 | 1.00 (Ref.) | - |
|  | T2 | 118/4465 | 1.17 (0.87~1.57) | 0.307 |
|  | T3 | 213/4371 | 1.35 (0.95~1.92) | 0.098 |
|  | *P* for trend | 0.097 | | |
| Relative cumulative PhenoAgeAccel | T1 | 136/4448 | 1.00 (Ref.) | - |
|  | T2 | 115/4468 | 1.06 (0.82~1.38) | 0.652 |
|  | T3 | 161/4423 | 1.54 (1.20~1.97) | 6.20×10^-4^ |
|  | *P* for trend | 4.66×10^-4^ | | |

Note: BioAgeAccel: biological aging acceleration; KDMAccel: Klemera-Doubal method biological age acceleration; PhenoAgeAccel: PhenoAge acceleration; T2D: type II diabetes; SD: standard deviation; T1: the first tertile; T2: the second tertile; T3: the third tertile; HR: hazard ratio; CI: confidence interval. Continuous (standardized) and categorized (tertiles) versions of BioAgeAccel and burdens were analyzed in separate models. All models were adjusted for sex, BMI, smoking status, drinking status, healthy diet score, physical activity, TDI, income, education, family history of T2D, and baseline BioAgeAccel.

Table S5. Association of BioAgeAccel burdens with the risk of all-cause mortality.

| BioAgeAccel burdens | | Events | HR (95%CI) | *P*-value |
| --- | --- | --- | --- | --- |
| KDMAccel slope | T1 | 200/4384 | 1.00 (Ref.) | - |
|  | T2 | 200/4384 | 1.22 (1.00~1.50) | 5.02×10^-2^ |
|  | T3 | 209/4375 | 1.47 (1.19~1.81) | 3.30×10^-4^ |
|  | *P* for trend | 3.26×10^-4^ | | |
| Cumulative KDMAccel | T1 | 196/4388 | 1.00 (Ref.) | - |
|  | T2 | 175/4408 | 0.87 (0.68~1.10) | 0232 |
|  | T3 | 238/4346 | 1.13 (0.83~1.54) | 0.448 |
|  | *P* for trend | 0.423 | | |
| Relative cumulative KDMAccel | T1 | 200/4384 | 1.00 (Ref.) | - |
|  | T2 | 179/4404 | 1.13 (0.92~1.39) | 0.232 |
|  | T3 | 230/4354 | 1.55 (1.26~1.90) | 2.77×10^-5^ |
|  | *P* for trend | 2.61×10^-5^ | | |
| PhenoAgeAccel slope | T1 | 134/4263 | 1.00 (Ref.) | - |
|  | T2 | 121/4275 | 1.07 (0.86~1.33) | 0.538 |
|  | T3 | 157/4240 | 1.72 (1.40~2.11) | 2.10×10^-7^ |
|  | *P* for trend | 1.04×10^-7^ | | |
| Cumulative PhenoAgeAccel | T1 | 79/4318 | 1.00 (Ref.) | - |
|  | T2 | 118/4278 | 1.00 (0.79~1.27) | 0.987 |
|  | T3 | 215/4182 | 1.21 (0.91~1.60) | 0.194 |
|  | *P* for trend | 0.172 | | |
| Relative cumulative PhenoAgeAccel | T1 | 136/4261 | 1.00 (Ref.) | - |
|  | T2 | 114/4282 | 1.03 (0.83~1.28) | 0.802 |
|  | T3 | 162/4235 | 1.75 (1.43~2.15) | 6.55×10^-8^ |
|  | *P* for trend | 2.34×10^-8^ | | |

Note: BioAgeAccel: biological aging acceleration; KDMAccel: Klemera-Doubal method biological age acceleration; PhenoAgeAccel: PhenoAge acceleration; T2D: type II diabetes; SD: standard deviation; T1: the first tertile; T2: the second tertile; T3: the third tertile; HR: hazard ratio; CI: confidence interval. Continuous (standardized) and categorized (tertiles) versions of BioAgeAccel and burdens were analyzed in separate models. All models were adjusted for sex, BMI, smoking status, drinking status, healthy diet score, physical activity, TDI, income, education, family history of T2D, and baseline BioAgeAccel.

Table S6. Association of BioAgeAccel transitions with incident T2D after excluding participants who developed T2D during the first two years of follow-up.

| BioAge | BioAgeAccel transitions | Events | HR (95%CI) | *P*-value |
| --- | --- | --- | --- | --- |
| KDM | Stable non-accelerated aging | 99/6604 | 1.00 (Ref.) | - |
|  | Non-accelerated to accelerated aging | 66/2833 | 1.54 (1.12~2.12) | 7.21×10^-3^ |
|  | Accelerated to non-accelerated aging | 36/905 | 2.00 (1.36~2.95) | 4.03×10^-4^ |
|  | Stable accelerated aging | 144/2932 | 2.46 (1.87~3.24) | 1.39×10^-10^ |
|  | Stable accelerated aging | 144/2932 | 1.00 (Ref.) | - |
|  | Accelerated to non-accelerated aging | 36/905 | 0.81 (0.56~1.18) | 0.278 |
| PhenoAge | Stable non-accelerated aging | 259/11710 | 1.00 (Ref.) | - |
|  | Non-accelerated to accelerated aging | 46/938 | 1.41 (1.02~1.94) | 0.038 |
|  | Accelerated to non-accelerated aging | 20/337 | 1.79 (1.13~2.84) | 0.013 |
|  | Stable accelerated aging | 20/289 | 1.53 (0.96~2.44) | 0.074 |
|  | Stable accelerated aging | 20/289 | 1.00 (Ref.) | - |
|  | Accelerated to non-accelerated aging | 20/337 | 1.17 (0.63~2.18) | 0.619 |

Note: BioAge: biological aging; BioAgeAccel: biological aging acceleration; KDM: Klemera-Doubal method biological age; T2D: type II diabetes; HR: hazard ratio; CI: confidence interval. Models were adjusted for age, sex, BMI, smoking status, drinking status, healthy diet score, physical activity, TDI, income, education, family history of T2D, and baseline BioAgeAccel.

Table S7. Association of BioAgeAccel burdens with incident T2D after excluding participants with less than two years of follow-up.

| BioAgeAccel burdens | | Events | HR (95%CI) | *P*-value |
| --- | --- | --- | --- | --- |
| KDMAccel slope | Per SD increment | 345/13274 | 1.17 (1.04~1.31) | 0.008 |
|  | T1 | 134/4406 | 1.00 (Ref.) | - |
|  | T2 | 94/4445 | 0.94 (0.71~1.23) | 0.634 |
|  | T3 | 117/4423 | 1.36 (1.04~1.78) | 0.027 |
|  | *P* for trend | 0.034 | | |
| Cumulative KDMAccel | Per SD increment | 345/13274 | 1.24 (1.01~1.52) | 0.041 |
|  | T1 | 69/4471 | 1.00 (Ref.) | - |
|  | T2 | 96/4443 | 0.97 (0.69~1.37) | 0.870 |
|  | T3 | 180/4360 | 1.14 (0.74~1.74) | 0.550 |
|  | *P* for trend | 0.489 | | |
| Relative cumulative KDMAccel | Per SD increment | 345/13274 | 1.13 (1.02~1.26) | 0.025 |
|  | T1 | 132/4408 | 1.00 (Ref.) | - |
|  | T2 | 89/4450 | 0.92 (0.70~1.22) | 0.571 |
|  | T3 | 124/4416 | 1.40 (1.08~1.83) | 0.012 |
|  | *P* for trend | 0.015 | | |
| PhenoAgeAccel slope | Per SD increment | 345/13274 | 1.19 (1.07~1.33) | 0.002 |
|  | T1 | 113/4427 | 1.00 (Ref.) | - |
|  | T2 | 103/4436 | 1.06 (0.80~1.40) | 0.705 |
|  | T3 | 129/4411 | 1.46 (1.11~1.91) | 0.007 |
|  | *P* for trend | 0.006 | | |
| Cumulative PhenoAgeAccel | Per SD increment | 345/13274 | 1.17 (0.96~1.41) | 0.112 |
|  | T1 | 75/4465 | 1.00 (Ref.) | - |
|  | T2 | 97/4442 | 1.06 (0.77~1.46) | 0.708 |
|  | T3 | 173/4367 | 1.22 (0.83~1.79) | 0.304 |
|  | *P* for trend | 0.295 | | |
| Relative cumulative PhenoAgeAccel | Per SD increment | 345/13274 | 1.21 (1.09~1.34) | 2.27×10^4^ |
|  | T1 | 115/4425 | 1.00 (Ref.) | - |
|  | T2 | 95/4444 | 0.99 (0.75~1.32) | 0.948 |
|  | T3 | 135/4405 | 1.45 (1.10~1.89) | 0.007 |
|  | *P* for trend | 0.006 | | |

Note: BioAgeAccel: biological aging acceleration; CVD: cardiovascular disease; SD: standard deviation; T1: the first tertile; T2: the second tertile; T3: the third tertile; HR: hazard ratio; CI: confidence interval. Continuous (standardized) and categorized (tertiles) versions of BioAgeAccel were analyzed in separate models. All models were adjusted for age, education, income, TDI, drinking status, smoking status, healthy diet score, physical activity, BMI, diabetes history, and family history of T2D.

Table S8. Association of BioAgeAccel transitions with all-cause mortality after excluding participants who died during the first two years of follow-up.

| BioAge | BioAgeAccel transitions | Events | HR (95%CI) | *P*-value |
| --- | --- | --- | --- | --- |
| KDM | Stable non-accelerated aging | 232/6487 | 1.00 (Ref.) | - |
|  | Non-accelerated to accelerated aging | 116/2799 | 1.34 (1.07~1.68) | 0.350 |
|  | Accelerated to non-accelerated aging | 41/907 | 1.17 (0.84~1.64) | 0.012 |
|  | Stable accelerated aging | 152/2949 | 1.67 (1.35~2.08) | 3.04×10^-6^ |
|  | Stable accelerated aging | 152/2949 | 1.00 (Ref.) | - |
|  | Accelerated to non-accelerated aging | 41/907 | 0.70 (0.50~0.99) | 0.045 |
| PhenoAge | Stable non-accelerated aging | 402/11609 | 1.00 (Ref.) | - |
|  | Non-accelerated to accelerated aging | 80/914 | 1.96 (1.54~2.50) | 6.39×10^-8^ |
|  | Accelerated to non-accelerated aging | 17/344 | 1.25 (0.77~2.04) | 0.368 |
|  | Stable accelerated aging | 42/275 | 3.01 (2.17~4.16) | 2.90×10^-11^ |
|  | Stable accelerated aging | 42/275 | 1.00 (Ref.) | - |
|  | Accelerated to non-accelerated aging | 17/344 | 0.42 (0.24~0.73) | 2.30×10^-3^ |

Note: BioAge: biological aging; BioAgeAccel: biological aging acceleration; KDM: Klemera-Doubal method biological age; T2D: type II diabetes; HR: hazard ratio; CI: confidence interval. Models were adjusted for age, sex, BMI, smoking status, drinking status, healthy diet score, physical activity, TDI, income, education, family history of T2D, and baseline BioAgeAccel.

Table S9. Association of BioAgeAccel burdens with all-cause mortality after excluding participants who died during the first two years of follow-up.

| BioAgeAccel burdens | | Events | HR (95%CI) | *P*-value |
| --- | --- | --- | --- | --- |
| KDMAccel slope | Per SD increment | 541/13142 | 1.17 (1.07~1.28) | 8.08×10^-4^ |
|  | T1 | 175/4386 | 1.00 (Ref.) | - |
|  | T2 | 179/4382 | 1.24 (1.00~1.53) | 0.052 |
|  | T3 | 187/4374 | 1.48 (1.19~1.85) | 5.32×10^-4^ |
|  | *P* for trend | 5.25×10^-4^ | | |
| Cumulative KDMAccel | Per SD increment | 541/13142 | 1.25 (1.06~1.49) | 0.010 |
|  | T1 | 176/4385 | 1.00 (Ref.) | - |
|  | T2 | 160/4401 | 0.88 (0.68~1.13) | 0.308 |
|  | T3 | 205/4356 | 1.07 (0.77~1.49) | 0.674 |
|  | *P* for trend | 0.657 | | |
| Relative cumulative KDMAccel | Per SD increment | 541/13142 | 1.14 (1.04~1.24) | 4.71×10^-3^ |
|  | T1 | 175/4386 | 1.00 (Ref.) | - |
|  | T2 | 159/4402 | 1.13 (0.91~1.41) | 0.264 |
|  | T3 | 207/4354 | 1.57 (1.27~1.95) | 4.19×10^-5^ |
|  | *P* for trend | 3.85×10^-5^ | | |
| PhenoAgeAccel slope | Per SD increment | 541/13142 | 1.24 (1.14~1.35) | 1.54×10^-6^ |
|  | T1 | 179/4382 | 1.00 (Ref.) | - |
|  | T2 | 158/4403 | 1.05 (0.84~1.32) | 0.647 |
|  | T3 | 204/4357 | 1.55 (1.25~1.93) | 8.53×10^-5^ |
|  | *P* for trend | 6.16×10^-5^ | | |
| Cumulative PhenoAgeAccel | Per SD increment | 541/13142 | 1.38 (1.18~1.61) | 4.31×10^-5^ |
|  | T1 | 122/4439 | 1.00 (Ref.) | - |
|  | T2 | 154/4407 | 1.00 (0.78~1.29) | 0.997 |
|  | T3 | 265/4296 | 1.14 (0.84~1.54) | 0.403 |
|  | *P* for trend | 0.381 | | |
| Relative cumulative PhenoAgeAccel | Per SD increment | 541/13142 | 1.26 (1.17~1.37) | 2.02×10^-8^ |
|  | T1 | 180/4381 | 1.00 (Ref.) | - |
|  | T2 | 147/4414 | 0.99 (0.79~1.24) | 0.915 |
|  | T3 | 214/4347 | 1.56 (1.26~1.93) | 5.64×10^-5^ |
|  | *P* for trend | 3.30×10^-5^ | | |

Note: BioAgeAccel: biological aging acceleration; CVD: cardiovascular disease; SD: standard deviation; T1: the first tertile; T2: the second tertile; T3: the third tertile; HR: hazard ratio; CI: confidence interval. Continuous (standardized) and categorized (tertiles) versions of BioAgeAccel were analyzed in separate models. All models were adjusted for age, education, income, TDI, drinking status, smoking status, healthy diet score, physical activity, BMI, diabetes history, and family history of T2D.

Table S10. Association of BioAgeAccel transitions with incident T2D after excluding participants who underwent drug treatments at baseline and the first follow-up.

| BioAge | BioAgeAccel transitions | Events | HR (95%CI) | *P*-value |
| --- | --- | --- | --- | --- |
| KDM | Stable non-accelerated aging | 108/6582 | 1.00 (Ref.) | - |
|  | Non-accelerated to accelerated aging | 74/2818 | 1.60 (1.19~2.16) | 2.07×10^-3^ |
|  | Accelerated to non-accelerated aging | 40/892 | 2.09 (1.45~3.01) | 8.01×10^-5^ |
|  | Stable accelerated aging | 155/2901 | 2.51 (1.93~3.27) | 7.32×10^-12^ |
|  | Stable accelerated aging | 155/2901 | 1.00 (Ref.) | - |
|  | Accelerated to non-accelerated aging | 40/892 | 0.83 (0.58~1.18) | 0.307 |
| PhenoAge | Stable non-accelerated aging | 276/11655 | 1.00 (Ref.) | - |
|  | Non-accelerated to accelerated aging | 52/939 | 1.49 (1.10~2.02) | 9.74×10^-3^ |
|  | Accelerated to non-accelerated aging | 24/325 | 2.09 (1.37~3.19) | 6.47×10^-4^ |
|  | Stable accelerated aging | 25/274 | 1.95 (1.28~2.96) | 1.89×10^-3^ |
|  | Stable accelerated aging | 25/274 | 1.00 (Ref.) | - |
|  | Accelerated to non-accelerated aging | 24/325 | 1.07 (0.61~1.89) | 0.804 |

Note: BioAgeAccel: biological aging acceleration; T2D: type II diabetes; HR: hazard ratio; CI: confidence interval. Models were adjusted for age, education, income, TDI, drinking status, smoking status, healthy diet score, physical activity, BMI, diabetes history, family history of T2D, and baseline BioAgeAccel.

Table S11. Association of BioAgeAccel burdens with incident T2D after excluding participants who underwent drug treatments at baseline and the first follow-up.

| BioAgeAccel burdens | | Events | HR (95%CI) | *P*-value |
| --- | --- | --- | --- | --- |
| KDMAccel slope | Per SD increment | 377/13193 | 1.16 (1.04~1.29) | 0.007 |
|  | T1 | 147/4377 | 1.00 (Ref.) | - |
|  | T2 | 101/4422 | 0.93 (0.72~1.21) | 0.604 |
|  | T3 | 129/4394 | 1.41 (1.09~1.83) | 0.008 |
|  | *P* for trend | 0.012 | | |
| Cumulative KDMAccel | Per SD increment | 377/13193 | 1.23 (1.01~1.50) | 0.040 |
|  | T1 | 73/4451 | 1.00 (Ref.) | - |
|  | T2 | 102/4421 | 1.00 (0.72~1.39) | 0.990 |
|  | T3 | 202/4321 | 1.26 (0.84~1.89) | 0.257 |
|  | *P* for trend | 0.204 | | |
| Relative cumulative KDMAccel | Per SD increment | 377/13193 | 1.13 (1.02~1.25) | 0.021 |
|  | T1 | 145/4379 | 1.00 (Ref.) | - |
|  | T2 | 99/4424 | 0.95 (0.73~1.23) | 0.697 |
|  | T3 | 133/4390 | 1.41 (1.09~1.81) | 0.008 |
|  | *P* for trend | 0.011 | | |
| PhenoAgeAccel slope | Per SD increment | 377/13193 | 1.24 (1.11~1.37) | 6.58×10^-5^ |
|  | T1 | 126/4398 | 1.00 (Ref.) | - |
|  | T2 | 108/4415 | 1.04 (0.80~1.36) | 0.769 |
|  | T3 | 143/4380 | 1.60 (1.23~2.08) | 4.14×10^-4^ |
|  | *P* for trend | 3.28×10^-4^ | | |
| Cumulative PhenoAgeAccel | Per SD increment | 377/13193 | 1.29 (1.07~1.55) | 0.008 |
|  | T1 | 73/4451 | 1.00 (Ref.) | - |
|  | T2 | 106/4417 | 1.13 (0.82~1.54) | 0.458 |
|  | T3 | 198/4325 | 1.28 (0.88~1.85) | 0.195 |
|  | *P* for trend | 0.192 | | |
| Relative cumulative PhenoAgeAccel | Per SD increment | 377/13193 | 1.25 (1.13~1.38) | 8.33×10^-6^ |
|  | T1 | 128/4396 | 1.00 (Ref.) | - |
|  | T2 | 102/4421 | 1.01 (0.77~1.33) | 0.933 |
|  | T3 | 147/4376 | 1.53 (1.18~1.98) | 0.001 |
|  | *P* for trend | 9.16×10^-4^ | | |

Note: BioAgeAccel: biological aging acceleration; KDMAccel: Klemera-Doubal method biological age acceleration; PhenoAgeAccel: PhenoAge acceleration; T2D: type II diabetes; SD: standard deviation; T1: the first tertile; T2: the second tertile; T3: the third tertile; HR: hazard ratio; CI: confidence interval. Continuous (standardized) and categorized (tertiles) versions of BioAgeAccel and burdens were analyzed in separate models. All models were adjusted for sex, BMI, smoking status, drinking status, healthy diet score, physical activity, TDI, income, education, family history of T2D, and baseline BioAgeAccel.

Table S12. Association of BioAgeAccel transitions with incident T2D: competing risk-adjusted analysis.

| BioAge | BioAgeAccel transitions | Events | HR (95%CI) | *P*-value |
| --- | --- | --- | --- | --- |
| KDM | Stable non-accelerated aging | 115/6390 | 1.00 (Ref.) | - |
|  | Non-accelerated to accelerated aging | 82/2727 | 1.68 (1.26~2.24) | 3.93×10^-4^ |
|  | Accelerated to non-accelerated aging | 43/869 | 2.09 (1.47~2.97) | 4.51×10^-5^ |
|  | Stable accelerated aging | 172/2792 | 2.58 (2.00~3.32) | 2.82×10^-13^ |
|  | Stable accelerated aging | 172/2792 | 1.00 (Ref.) | - |
|  | Accelerated to non-accelerated aging | 43/869 | 0.81 (0.58~1.14) | 0.225 |
| PhenoAge | Stable non-accelerated aging | 302/11334 | 1.00 (Ref.) | - |
|  | Non-accelerated to accelerated aging | 57/867 | 1.53 (1.14~2.05) | 4.08×10^-3^ |
|  | Accelerated to non-accelerated aging | 24/324 | 1.83 (1.20~2.79) | 5.10×10^-3^ |
|  | Stable accelerated aging | 29/253 | 2.04 (1.38~3.02) | 3.73×10^-4^ |
|  | Stable accelerated aging | 29/253 | 1.00 (Ref.) | - |
|  | Accelerated to non-accelerated aging | 24/324 | 0.90 (0.52~1.54) | 0.693 |

Note: BioAgeAccel: biological aging acceleration; T2D: type II diabetes; HR: hazard ratio; CI: confidence interval. Models were adjusted for age, education, income, TDI, drinking status, smoking status, healthy diet score, physical activity, BMI, diabetes history, family history of T2D, and baseline BioAgeAccel.

Table S13. Association of BioAgeAccel burdens with incident T2D: competing risk-adjusted analysis.

| BioAgeAccel burdens | | Events | HR (95%CI) | *P*-value |
| --- | --- | --- | --- | --- |
| KDMAccel slope | Per SD increment | 412/12778 | 1.20 (1.08~1.33) | 7.41×10^-4^ |
|  | T1 | 158/4239 | 1.00 (Ref.) | - |
|  | T2 | 108/4288 | 0.94 (0.73~1.21) | 0.651 |
|  | T3 | 146/4251 | 1.51 (1.18~1.94) | 9.84×10^-4^ |
|  | *P* for trend | 0.001 | | |
| Cumulative KDMAccel | Per SD increment | 412/12778 | 1.27 (1.05~1.53) | 0.001 |
|  | T1 | 79/4318 | 1.00 (Ref.) | - |
|  | T2 | 110/4286 | 0.99 (0.72~1.36) | 0.946 |
|  | T3 | 223/4174 | 1.28 (0.87~1.90) | 0.212 |
|  | *P* for trend | 0.158 | | |
| Relative cumulative KDMAccel | Per SD increment | 412/12778 | 1.15 (1.04~1.27) | 6.33×10^-3^ |
|  | T1 | 156/4241 | 1.00 (Ref.) | - |
|  | T2 | 106/4290 | 0.96 (0.75~1.24) | 0.757 |
|  | T3 | 150/4247 | 1.50 (1.17~1.91) | 1.14×10^-3^ |
|  | *P* for trend | 0.001 | | |
| PhenoAgeAccel slope | Per SD increment | 412/12778 | 1.26 (1.15~1.39) | 3.14×10^-6^ |
|  | T1 | 134/4263 | 1.00 (Ref.) | - |
|  | T2 | 121/4275 | 1.11 (0.86~1.44) | 0.415 |
|  | T3 | 157/4240 | 1.61 (1.25~2.07) | 1.91×10^-4^ |
|  | *P* for trend | 1.46×10^-4^ | | |
| Cumulative PhenoAgeAccel | Per SD increment | 412/12778 | 1.28 (1.08~1.51) | 4.92×10^-3^ |
|  | T1 | 79/4318 | 1.00 (Ref.) | - |
|  | T2 | 118/4278 | 1.18 (0.87~1.59) | 0.290 |
|  | T3 | 215/4182 | 1.32 (0.93~1.89) | 0.125 |
|  | *P* for trend | 0.127 | | |
| Relative cumulative PhenoAgeAccel | Per SD increment | 412/12778 | 1.24 (1.13~1.36) | 4.73×10^-6^ |
|  | T1 | 136/4261 | 1.00 (Ref.) | - |
|  | T2 | 114/4282 | 1.07 (0.82~1.39) | 0.611 |
|  | T3 | 162/4235 | 1.58 (1.23~2.02) | 3.01×10^-4^ |
|  | *P* for trend | 2.18×10^-4^ | | |

Note: BioAgeAccel: biological aging acceleration; KDMAccel: Klemera-Doubal method biological age acceleration; PhenoAgeAccel: PhenoAge acceleration; T2D: type II diabetes; SD: standard deviation; T1: the first tertile; T2: the second tertile; T3: the third tertile; HR: hazard ratio; CI: confidence interval. Continuous (standardized) and categorized (tertiles) versions of BioAgeAccel and burdens were analyzed in separate models. All models were adjusted for sex, BMI, smoking status, drinking status, healthy diet score, physical activity, TDI, income, education, family history of T2D, and baseline BioAgeAccel.

Table S14. Scoring of the Finnish Diabetes Risk Score (FINDRISC).

| Points | Age | Family history of diabetes | Daily fruit/veg. | Physical activity | Blood pressure medication | History of high blood glucose | body mass index (kg/m^2^) | Waist circumference (cm) | |
| --- | --- | --- | --- | --- | --- | --- | --- | --- | --- |
|  |  |  |  |  |  |  |  | Women | Men |
| 0 | <45 | No | Yes | Yes | No | No | ≤25 | <80 | <94 |
| 1 |  |  | No |  |  |  | 25~30 |  |  |
| 2 | 45~54 |  |  | No | Yes |  |  |  |  |
| 3 | 55~64 |  |  |  |  |  | >30 | 80~88 | 94~102 |
| 4 | ≥65 |  |  |  |  |  |  | ≥88 | ≥102 |
| 5 |  | Yes |  |  |  | Yes |  |  |  |

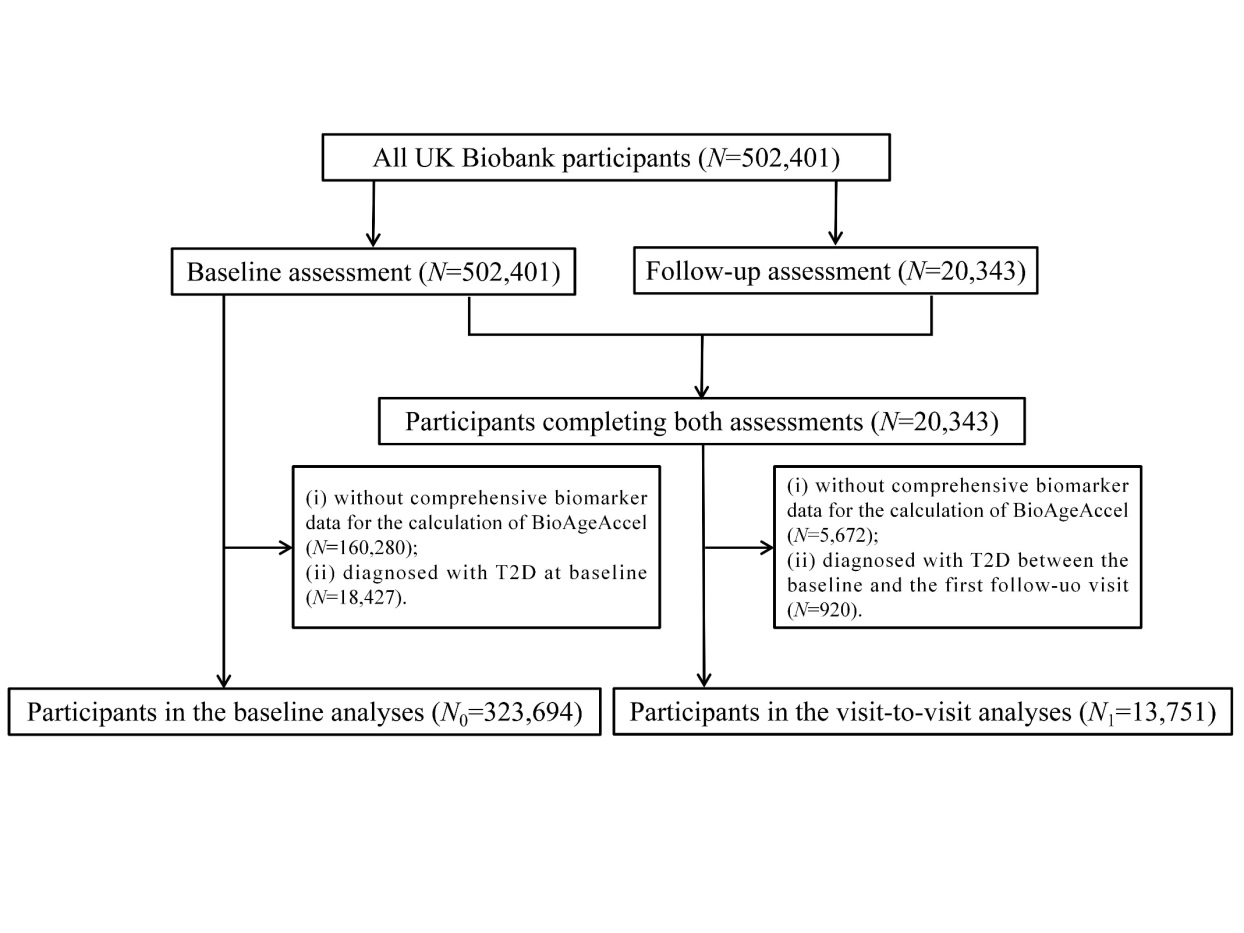

Figure S1. Flowchart of the process of sample selection. BioAgeAccel: biological aging acceleration. T2D: type II diabetes.

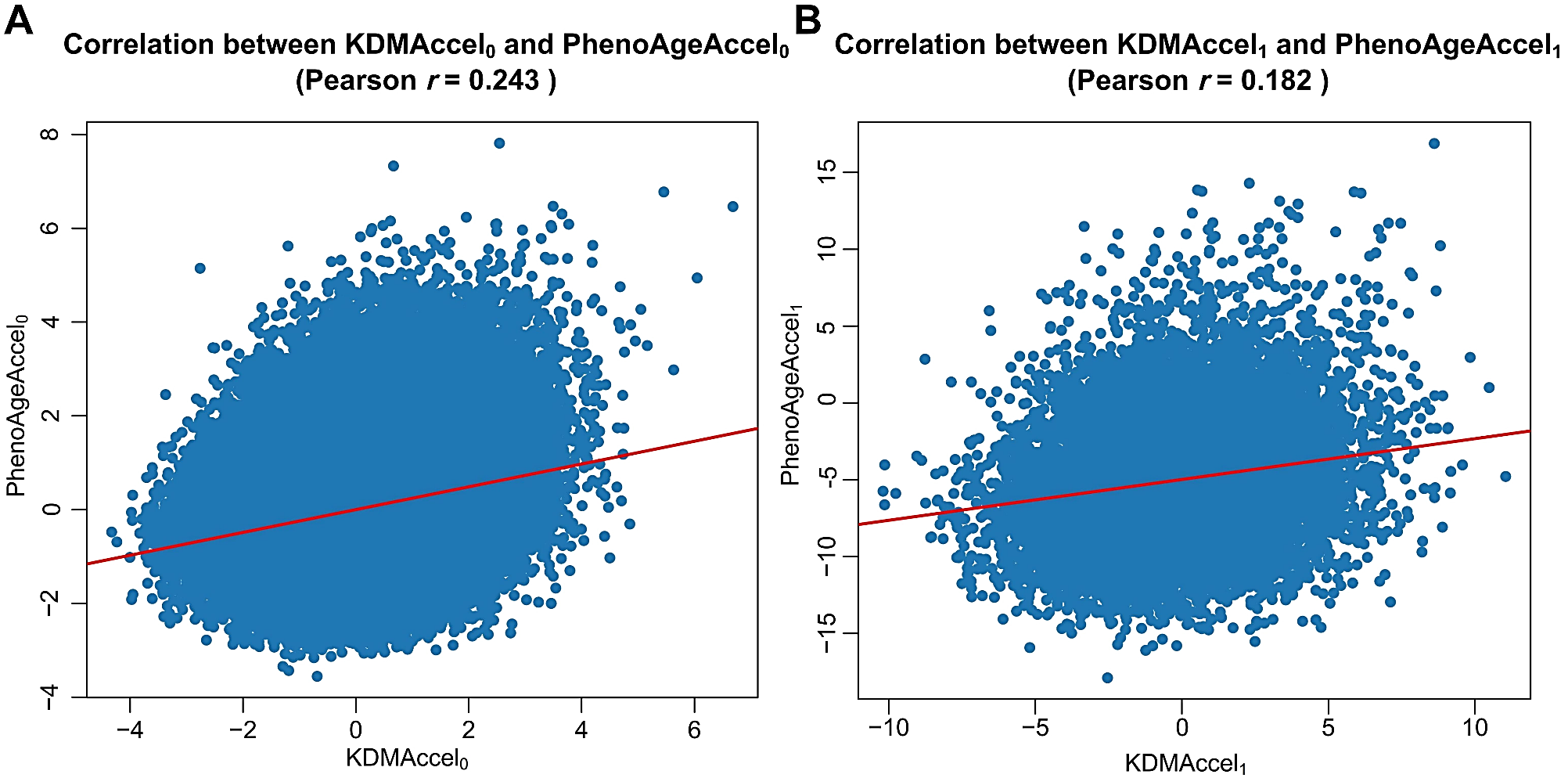

Figure S2. Correlation between KDMAccel and PhenoAgeAccel. (A) Baseline biological age acceleration (KDMAccel_0_ and PhenoAgeAccel_0_); (B) Longitudinal change in BioAgeAccel (KDMAccel_1_ and PhenoAgeAccel_1_) between visits. Each point represents an individual participant, with the red line indicating linear regression fit. BioAgeAccel: biological aging acceleration; KDMAccel: Klemera-Doubal method biological age acceleration; PhenoAgeAccel: PhenoAge acceleration. KDMAccel_0_ and KDMAccel_1_ referred to KDMAccel for each participant at baseline and the first follow-up, respectively. PhenoAgeAccel_0_ and PhenoAgeAccel_1_ referred to PhenoAgeAccel for each participant at baseline and the first follow-up, respectively.

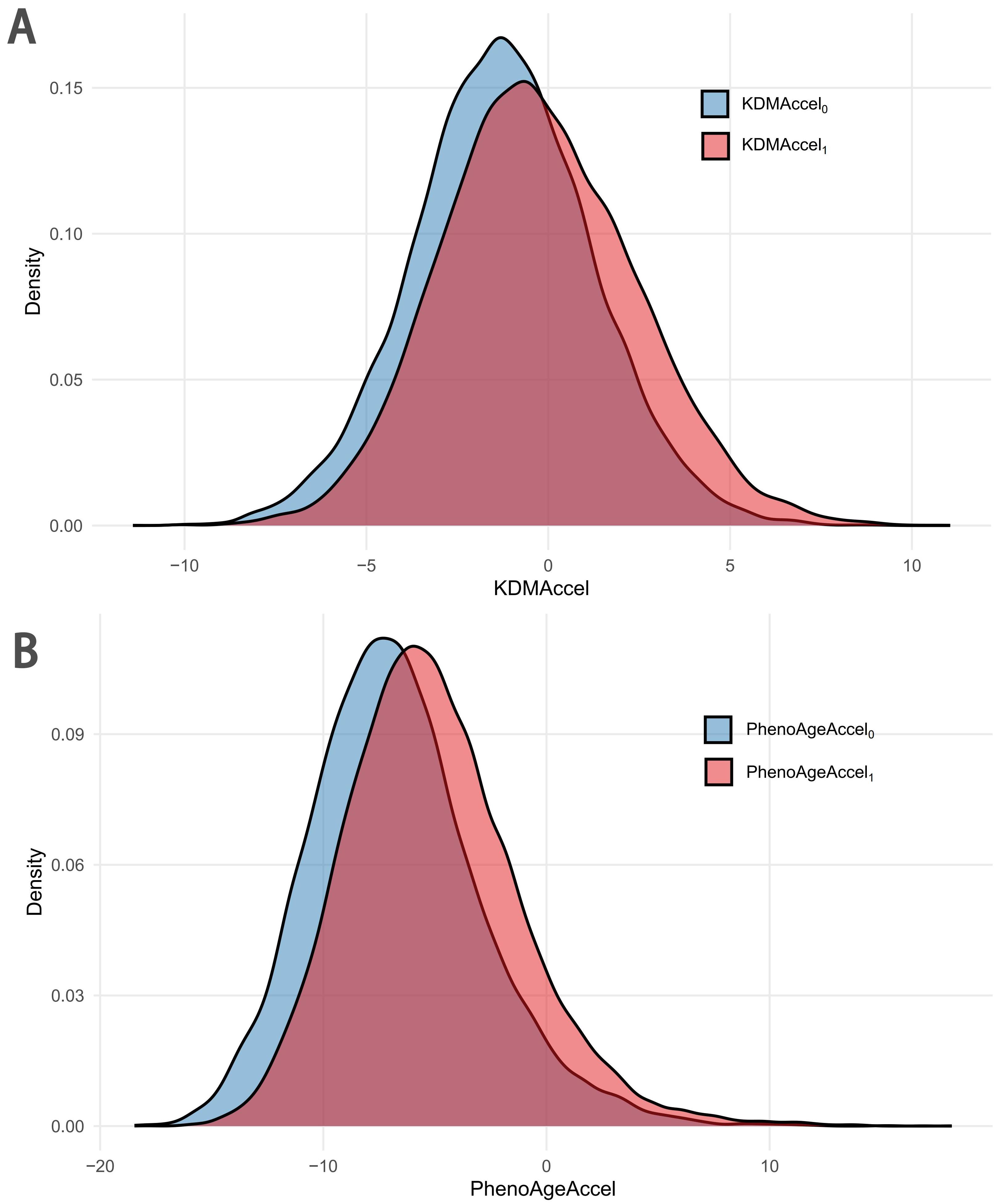

Figure S3. Distribution of biological age acceleration measures at baseline and follow-up. (A) Density plots of the distribution of KDMAccel_0_ and KDMAccel_1_ among populations across two assessments; (B) Density plots of the distribution of PhenoAgeAccel_0_ and PhenoAgeAccel_1_ among populations across two assessments. BioAgeAccel: biological aging acceleration; KDMAccel: Klemera-Doubal method biological age acceleration; PhenoAgeAccel: PhenoAge acceleration. KDMAccel_0_ and KDMAccel_1_ referred to KDMAccel for each participant at baseline and the first follow-up, respectively. PhenoAgeAccel_0_ and PhenoAgeAccel_1_ referred to PhenoAgeAccel for each participant at baseline and the first follow-up, respectively.

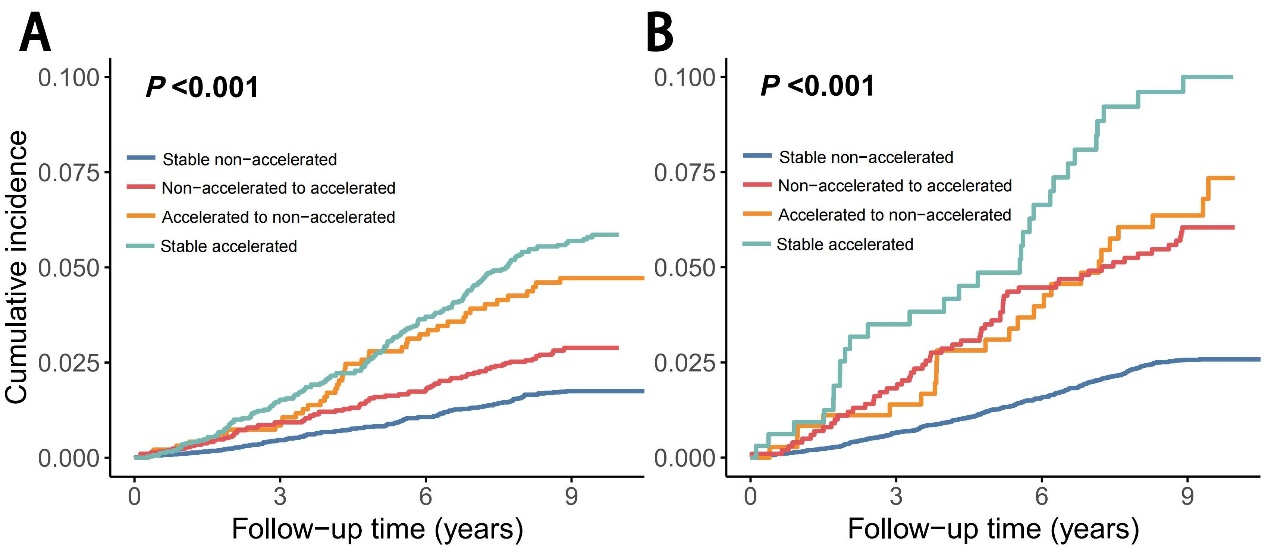

Figure S4. Cumulative incidence of T2D by BioAgeAccel transition status. (A) Kaplan-Meier curves of cumulative incidence of T2D events stratified by KDMAccel transitions; (B) Kaplan-Meier curves of cumulative incidence of T2D events stratified by PhenoAgeAccel transitions. BioAgeAccel: biological aging acceleration; T2D: type II diabetes.

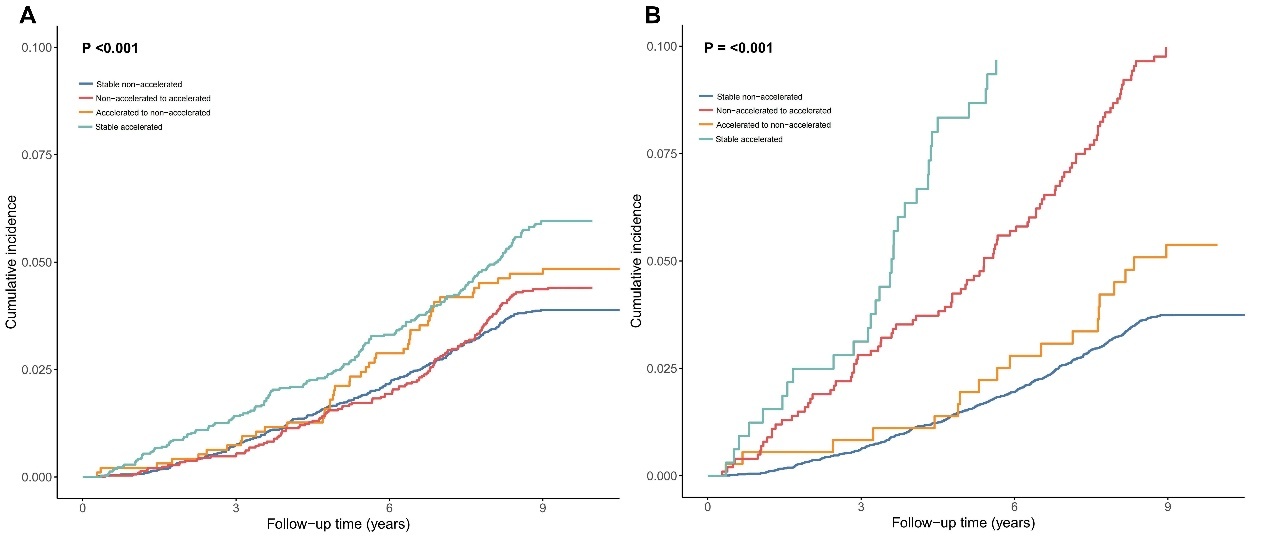

Figure S4. Cumulative incidence of all-cause mortality by BioAgeAccel transition status. (A) Kaplan-Meier curves of cumulative incidence of all-cause mortality events stratified by KDMAccel transitions; (B) Kaplan-Meier curves of cumulative incidence of all-cause mortality events stratified by PhenoAgeAccel transitions. BioAgeAccel: biological aging acceleration.

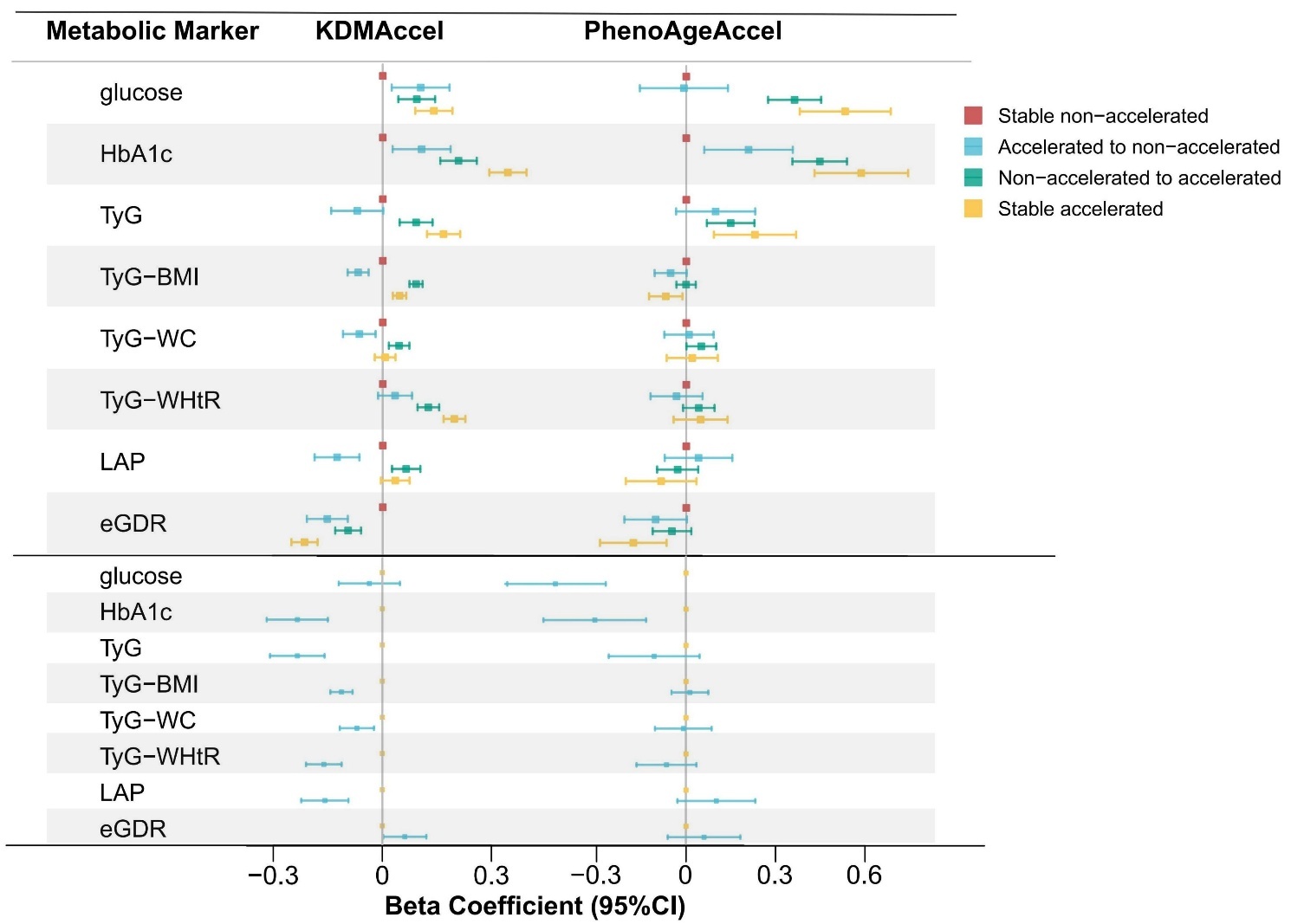

Figure S6. Forest plots of the association of BioAgeAccel transitions with glycemic traits in the visit-to-visit analyses. BioAgeAccel: biological aging acceleration; KDMAccel: Klemera-Doubal method biological age acceleration; PhenoAgeAccel: phenotypic age acceleration; T2D: type II diabetes; TyG: triglyceride-glucose; BMI: body mass index; WC: waist circumference; WHtR: waist-to-height ratio; LAP: lipid accumulation product; eGDR: estimated glucose disposal rate.

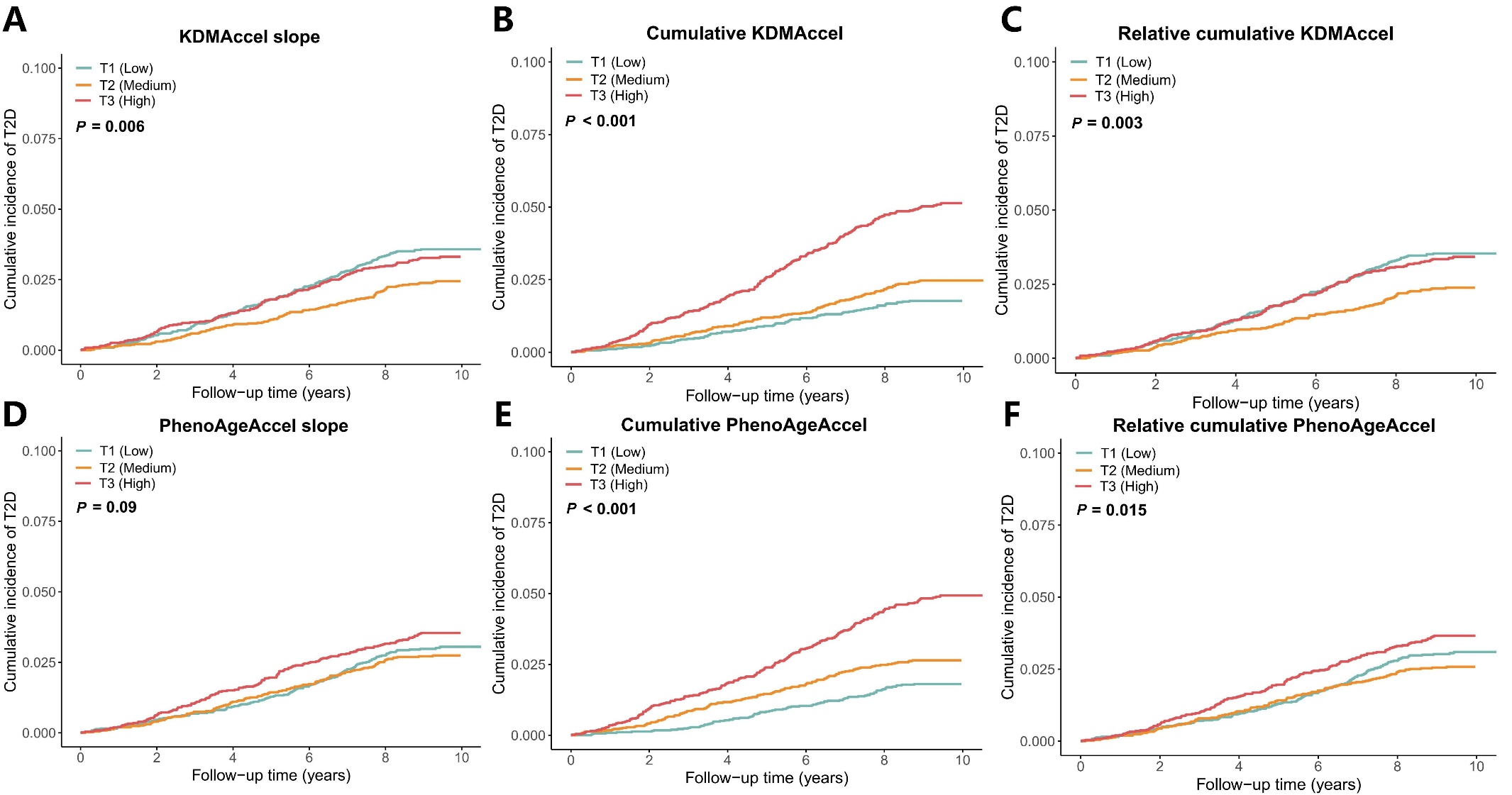

Figure S7. Cumulative incidence of T2D by BioAgeAccel burdens status. Kaplan-Meier curves of cumulative incidence of T2D events stratified by (A) KDMAccel slope; (B) Cumulative KDMAccel; (C) Relative cumulative KDMAccel; (D) PhenoAgeAccel slope; (E) Cumulative PhenoAgeAccel; (F) Relative cumulative PhenoAgeAccel. BioAgeAccel: biological aging acceleration; KDMAccel: Klemera-Doubal method biological age acceleration; PhenoAgeAccel: PhenoAge acceleration; T2D: type II diabetes.

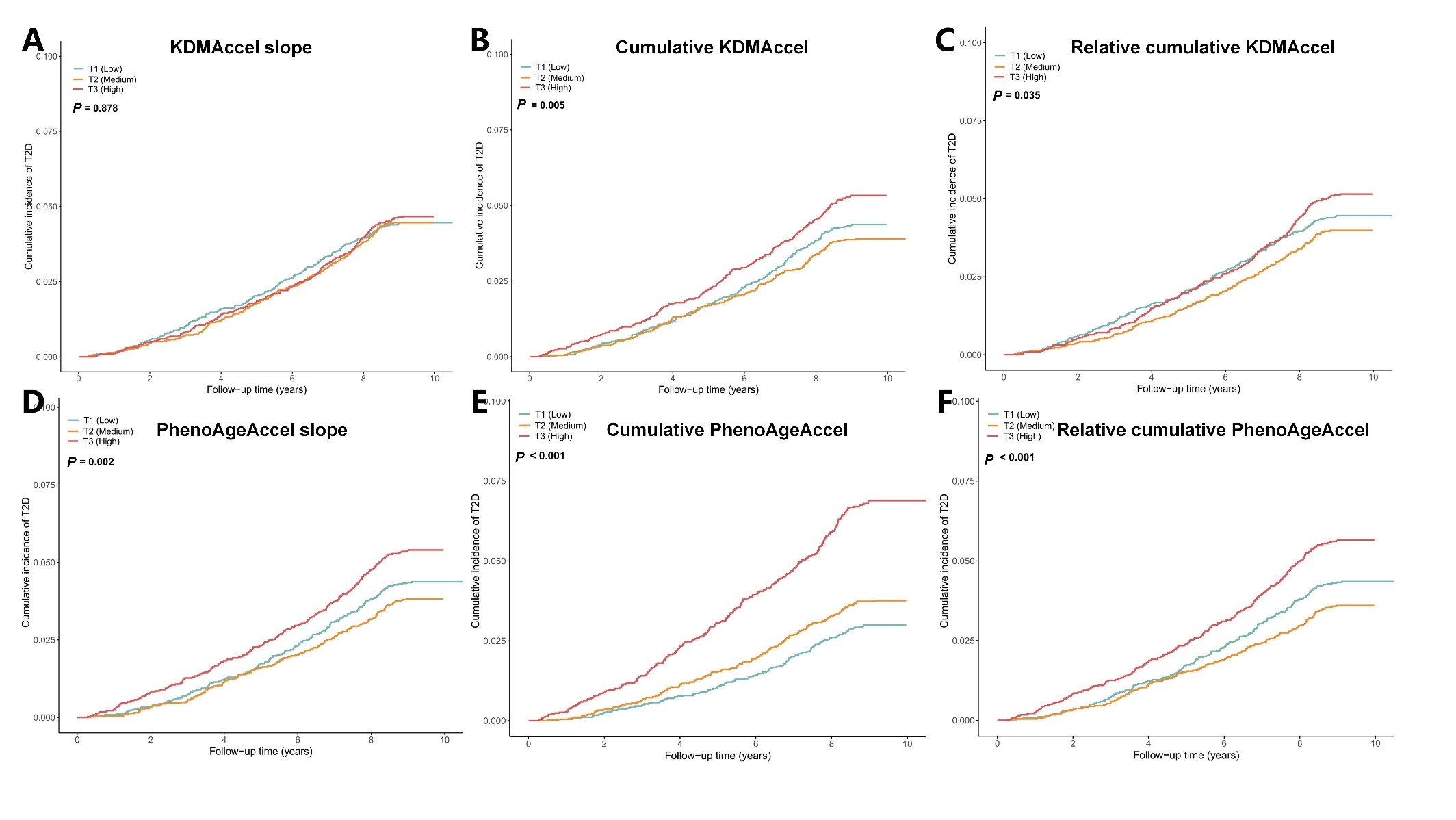

Figure S6. Cumulative incidence of all-cause mortality by BioAgeAccel burdens status. Kaplan-Meier curves of cumulative incidence of all-cause mortality events stratified by (A) KDMAccel slope; (B) Cumulative KDMAccel; (C) Relative cumulative KDMAccel; (D) PhenoAgeAccel slope; (E) Cumulative PhenoAgeAccel; (F) Relative cumulative PhenoAgeAccel. BioAgeAccel: biological aging acceleration; KDMAccel: Klemera-Doubal method biological age acceleration; PhenoAgeAccel: PhenoAge acceleration.

**References**

1. Er, L.K.*, et al.* (2016) Triglyceride Glucose-Body Mass Index Is a Simple and Clinically Useful Surrogate Marker for Insulin Resistance in Nondiabetic Individuals, *PloS one*, **11**, e0149731.
2. Kahn, H.S. (2005) The "lipid accumulation product" performs better than the body mass index for recognizing cardiovascular risk: a population-based comparison, *BMC cardiovascular disorders*, **5**, 26.
3. Kwon, D. and Belsky, D.W. (2021) A toolkit for quantification of biological age from blood chemistry and organ function test data: BioAge, *Geroscience*, **43**, 2795-2808.
4. Levine, M.E.*, et al.* (2018) An epigenetic biomarker of aging for lifespan and healthspan, *Aging*, **10**, 573-591.
5. Lim, J.*, et al.* (2019) Comparison of triglyceride glucose index, and related parameters to predict insulin resistance in Korean adults: An analysis of the 2007-2010 Korean National Health and Nutrition Examination Survey, *PloS one*, **14**, e0212963.
6. Liu, Z.*, et al.* (2018) A new aging measure captures morbidity and mortality risk across diverse subpopulations from NHANES IV: A cohort study, *PLoS medicine*, **15**, e1002718.
7. Mak, J.K.L.*, et al.* (2023) Clinical biomarker-based biological aging and risk of cancer in the UK Biobank, *Br. J. Cancer*, **129**, 94-103.
8. Ramdas Nayak, V.K.*, et al.* (2022) Triglyceride Glucose (TyG) Index: A surrogate biomarker of insulin resistance, *JPMA. The Journal of the Pakistan Medical Association*, **72**, 986-988.
9. van Buuren, S. and Groothuis-Oudshoorn, K. (2011) mice: Multivariate Imputation by Chained Equations in R, *J. Stat. Softw.*, **45**, 1-67.
10. Zhang, Z.*, et al.* (2024) Insulin resistance assessed by estimated glucose disposal rate and risk of incident cardiovascular diseases among individuals without diabetes: findings from a nationwide, population based, prospective cohort study, *Cardiovasc. Diabetol.*, **23**, 194.
11. Zheng, S.*, et al.* (2016) Triglyceride glucose-waist circumference, a novel and effective predictor of diabetes in first-degree relatives of type 2 diabetes patients: cross-sectional and prospective cohort study, *Journal of translational medicine*, **14**, 260.
